# Multi-ancestry Genome-wide Association Study of Varicose Veins Reveals Polygenic Architecture, Genetic Overlap with Arterial and Venous Disease, and Novel Therapeutic Opportunities

**DOI:** 10.1101/2022.02.22.22271350

**Authors:** Michael G. Levin, Jennifer E. Huffman, Anurag Verma, Kyle A. Sullivan, Alexis A. Rodriguez, David Kainer, Michael R. Garvin, Matthew Lane, Hyejung Won, Binglan Li, Yuan Luo, Gail P. Jarvik, Hakon Hakonarson, Elizabeth A. Jasper, Alexander G. Bick, Marylyn D. Ritchie, Daniel A. Jacobson, Ravi K. Madduri, Scott M. Damrauer, the VA Million Veteran Program

## Abstract

**Background:** Varicose veins represent a common cause of cardiovascular morbidity, with limited available medical therapies. Although varicose veins are heritable and epidemiologic studies have identified several candidate varicose veins risk factors, the molecular and genetic basis remains uncertain. Here, we analyzed the contribution of common genetic variants to varicose veins using data from the VA Million Veteran Program and other large multi-ancestry biobanks. Among 49,765 individuals with varicose veins and 1,334,301 disease-free controls, we identified 139 risk loci. We identified genetic overlap between varicose veins, other vascular diseases, and dozens of anthropometric factors. Using Mendelian randomization, we prioritized novel therapeutic targets via integration of proteomic and transcriptomic data. Finally, topological enrichment analyses confirmed the biologic roles of endothelial shear flow disruption, inflammation, vascular remodeling, and angiogenesis. These findings may facilitate future efforts to develop non-surgical therapies for varicose veins.

## INTRODUCTION

Varicose veins are a manifestation of chronic venous disease, characterized by venous dilation resulting from venous valvular incompetence, thrombosis, or non-thrombotic venous occlusion. While population prevalence varies, increasing with age and BMI, some estimates suggest varicose veins affect nearly 40% of the adult population.^1–8^ Varicose veins may initially present asymptomatically as superficial dilation and tortuosity, but may progress to include symptoms of pain, edema, skin changes, and ulceration.^9^ Serious downstream cardiovascular consequences have been ascribed to varicose veins, including risks of venous thromboembolism and peripheral artery disease.^10,11^

Several molecular mechanisms have been implicated in varicose veins.^12–15^ The generally accepted molecular pathology of varicose veins involves five major processes: (1) disrupted vascular endothelium homeostasis due to altered shear stress from increases in venous pressure and/or valve failure resulting in (2) immune cell activation, adhesion, and infiltration of the blood vessel wall causing (3) local inflammatory processes and cytokine production followed by (4) remodeling of the extracellular matrix and (5) angiogenesis.

Observational studies have identified several risk factors for varicose veins, including age, sex, measures of obesity (eg. body mass index, waist-to-hip ratio), height, prior venous thromboembolism, pregnancy, and family history of varicose veins, among others.^16,17^ Recent genome wide association studies (GWAS) have begun to elucidate common genetic risk factors for varicose veins, identifying more than 30 risk loci.^16,18,19^ However, these studies have been limited to individuals of European ancestry, and a substantial proportion of varicose veins heritability remains unexplained.

Here, we investigated the genetic basis of varicose veins by conducting the largest multi-ancestry GWAS of varicose veins to date. Using downstream bioinformatic techniques we aimed to (1) prioritize causal risk variants, (2) identify traits with shared genetic mechanisms, (3) define putative causal risk factors, (4) identify circulating proteins and drug targets that may represent therapeutic targets, and (5) provide mechanistic understanding of varicose veins biology using a multi-omic integration method.

## RESULTS

### Multi-ancestry GWAS

Overall GWAS and study design are presented in Error! Reference source not found.. The VA Million Veteran Program (MVP) is a national research program established in 2011, designed to understand how genes, lifestyle, and military exposures affect health and illness.^20^ Using electronic health record diagnosis codes (**SUPPLEMENTAL METHODS**), we identified 18,977 MVP participants with varicose veins and 561,203 disease-free individuals. Leveraging the diverse genetic composition of MVP, we performed ancestry-specific GWAS. Next, we assembled genetic association data for 49,765 individuals with varicose veins and 1,334,301 disease-free controls, across four major genetic ancestral populations (African, East-Asian, European, and Hispanic) by supplementing our MVP analysis with summary data from other large-scale biobanks (eMERGE, UK Biobank, FinnGen, and BioBank Japan). We combined genetic association data first within, and then across ancestries, using fixed-effects meta-analysis.

Following multi-ancestry meta-analysis, we identified 8,336 DNA sequence variants at 139 loci (r^2^ < 0.1, distance = 250kb) associated with varicose veins at genome-wide significance (p < 5 × 10^−8^)(Error! Reference source not found.Error! Reference source not found.; **SUPPLEMENTAL FIGURE 1; SUPPLEMENTAL TABLES 1-2**). We replicated 30/36 previously-reported risk loci^16,18,19^ (27/36 loci at genome-wide significance, and 3/9 remaining loci at nominal significance [p < 0.05]; **SUPPLEMENTAL TABLE 3**). Overall, novel associations included variants with lower frequency and of greater effect size than previously reported associations, including 11 rare variants (minor allele frequency < 0.01) (**SUPPLEMENTAL FIGURE 2**). Using linkage disequilibrium score regression^21^, we found the majority of genomic inflation was due to polygenicity rather than population stratification (LD score intercept = 1.08 [0.01]; Ratio: 0.14 [0.02]). We estimate the heritability of varicose veins at 15.7% (95% Confidence Interval [CI] 13.6% to 17.8%), based on an estimated population prevalence of 23%.^22^

### Fine-mapping of risk loci

To refine the set of genetic variants associated with varicose veins, we leveraged the multi-ancestry nature of our meta-analysis to perform multi-ancestry fine-mapping using MR-MEGA.^23^ This statistical approach performs meta-regression accounting for the heterogeneity in allelic effects that is correlated with ancestry to construct credible sets. The median size of the 99% credible sets was 1 variant, while the mean size was 36. We identified 70 varicose veins risk loci where a single variant was included in the 99% credible set (**SUPPLEMENTAL TABLE 4**). At 63 of these 70 loci, multi-ancestry fine-mapping prioritized a different variant than the fixed-effects meta-analysis. Novel risk variants included rs6025, which represents the thrombophilic mutation F5 p.Arg534Gln (factor V Leiden). This association reinforces a pathophysiologic role for venous thromboembolism, which is a clinically-recognized secondary etiology of varicose veins within the CEAP (Clinical-Etiology-Anatomy-Pathophysiology) classification of chronic venous disorders.^9^ Similarly, rs62033413 at the *FTO* locus (associated with increased weight, body mass index, and hip circumference, among others), highlights an association between obesity and risk of varicose veins.

### Tissue and Gene-set Enrichment

We evaluated whether specific tissues were enriched for varicose veins GWAS loci using MAGMA, which integrates GWAS summary statistics with gene expression (RNAseq) data from the Genotype Tissue Expression Project (GTEx v8). We identified significant enrichment in vascular tissues including the aorta, tibial and coronary arteries; reproductive tissues including uterus, ovary, cervix (endo-and ectocervix), and breast; as well as tibial nerve, subcutaneous adipose, and sigmoid colon (**SUPPLEMENTAL FIGURE 3A**). MAGMA gene-set enrichment prioritized significant (Bonferroni adjusted p < 0.05) roles for vascular smooth muscle cell differentiation, skeletal system morphogenesis, and Wnt signaling, among others (**SUPPLEMENTAL FIGURE 3B**).

As an alternative to the polygenic mapping approach utilized by MAGMA, we also tested whether genes located nearest to genome-wide significant loci were associated with specific biological processes. Here, we found significant (FDR < 0.05) enrichment for pathways involving vascular smooth muscle cell differentiation (based on associations near *PDGFB, NFATC2, NFATC1*, and *NOTCH1*), as well as pathways related to vascular, cardiac, and muscle growth/development/differentiation (**SUPPLEMENTAL TABLE 5**). Of 24 genes previously implicated in Mendelian diseases associated with varicose veins in the Human Phenotype Ontology database (**SUPPLEMENTAL TABLE 6**), only 1 (PIEZO1) was the closest gene to a genome-wide significant GWAS locus (**SUPPLEMENTAL TABLE 2**).

### Shared genetic architecture of vascular traits

Given the polygenic architecture of varicose veins and other vascular traits, as well as observational associations linking varicose veins with other vascular diseases, we estimated genetic correlations between varicose veins and other common vascular diseases. Varicose veins were significantly (FDR q < 0.05) genetically correlated with both venous (venous thromboembolism r_g_ = 0.27, p = 4.8 × 10^−9^) and arterial traits (abdominal aortic aneurysm r_g_ = 0.1, p = 2.1 × 10^−4^; peripheral artery disease r_g_ = 0.21, p = 8.6 × 10^−8^), although the degree of correlation was modest (**SUPPLEMENTAL FIGURE 4**).

### Phenome-wide scan and colocalization reveal pleiotropic associations

To characterize the broader spectrum of traits sharing risk variants with varicose veins, we performed a phenome-wide association study (pheWAS) followed by colocalization. We queried associations among GWAS of diagnoses/operations/medications/surveys/measurements (n = 2514) among UK Biobank participants, GWAS of circulating proteins (n = 4489), and GWAS of circulating metabolites (n = 974) from the MRC IEU Open GWAS Project. Among categories of UK Biobank traits sharing genome-wide significant associations (p < 5 × 10^−8^) with sentinel varicose veins SNPs were 81 anthropometric traits, 20 diagnoses, 15 operations, 5 medications/treatments, and 71 other survey/self-reported traits (**FIGURE 2**). We further identified 77 circulating proteins, and 216 circulating metabolites that shared associations with sentinel varicose vein SNPs (**FIGURE 2**).

**FIGURE 1:**
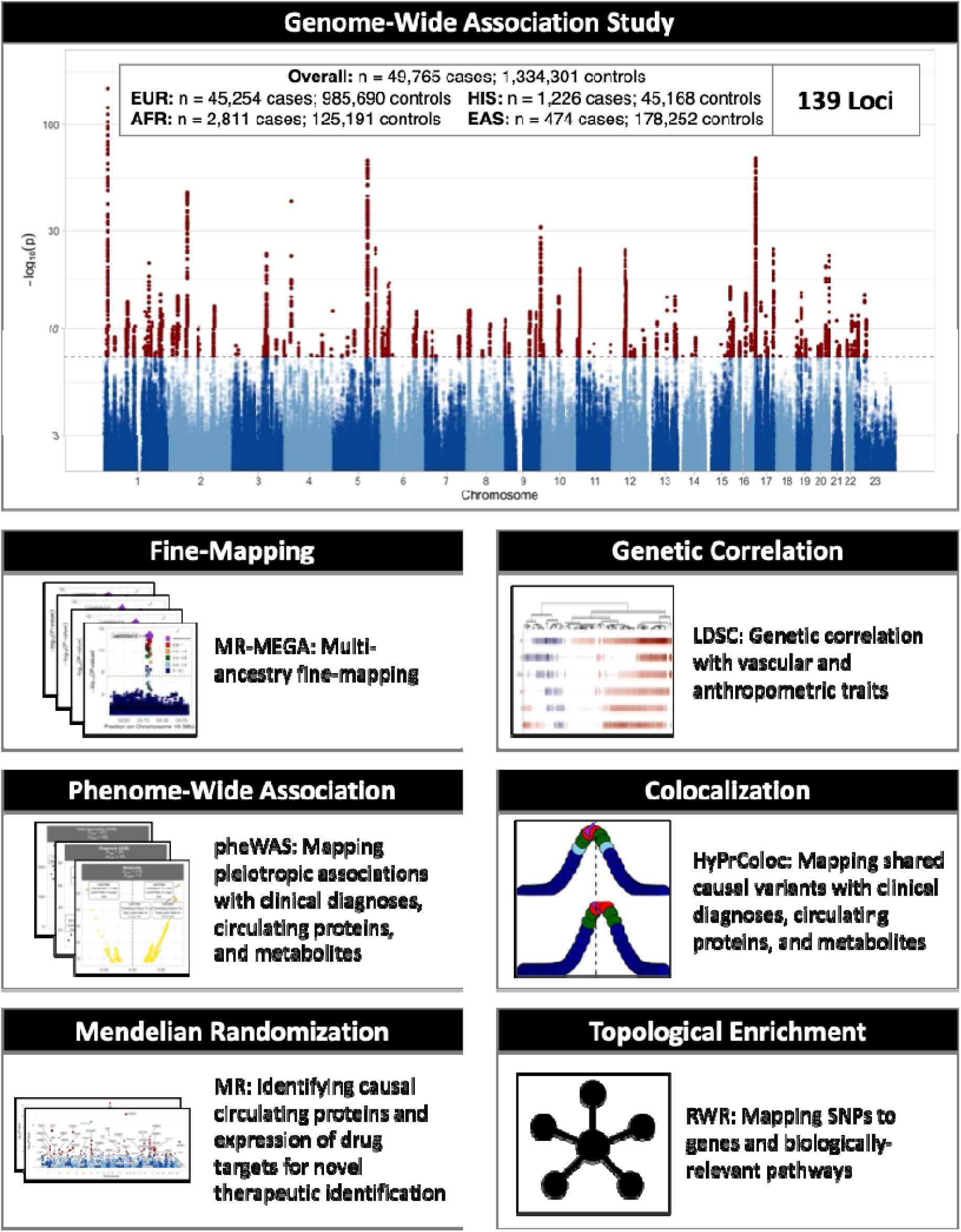
Study Design. Overview of study design. A multi-ancestry genome wide association study meta-analysis is highlighted in the top portion of the figure. Downstream bioinformatic analyses were then performed to identify causal loci, characterize genetic relationships with other traits/diseases, understand novel therapeutic opportunities, and implicate biologically-relevant pathways. EUR = European-ancestry, AFR = African-ancestry, HIS = Hispanic, EAS = East-Asian ancestry, LDSC = linkage disequilibrium score regression, PheWAS = phenome-wide association study, MR = Mendelian randomization, RWR = random-walk with restart.

**FIGURE 2:**
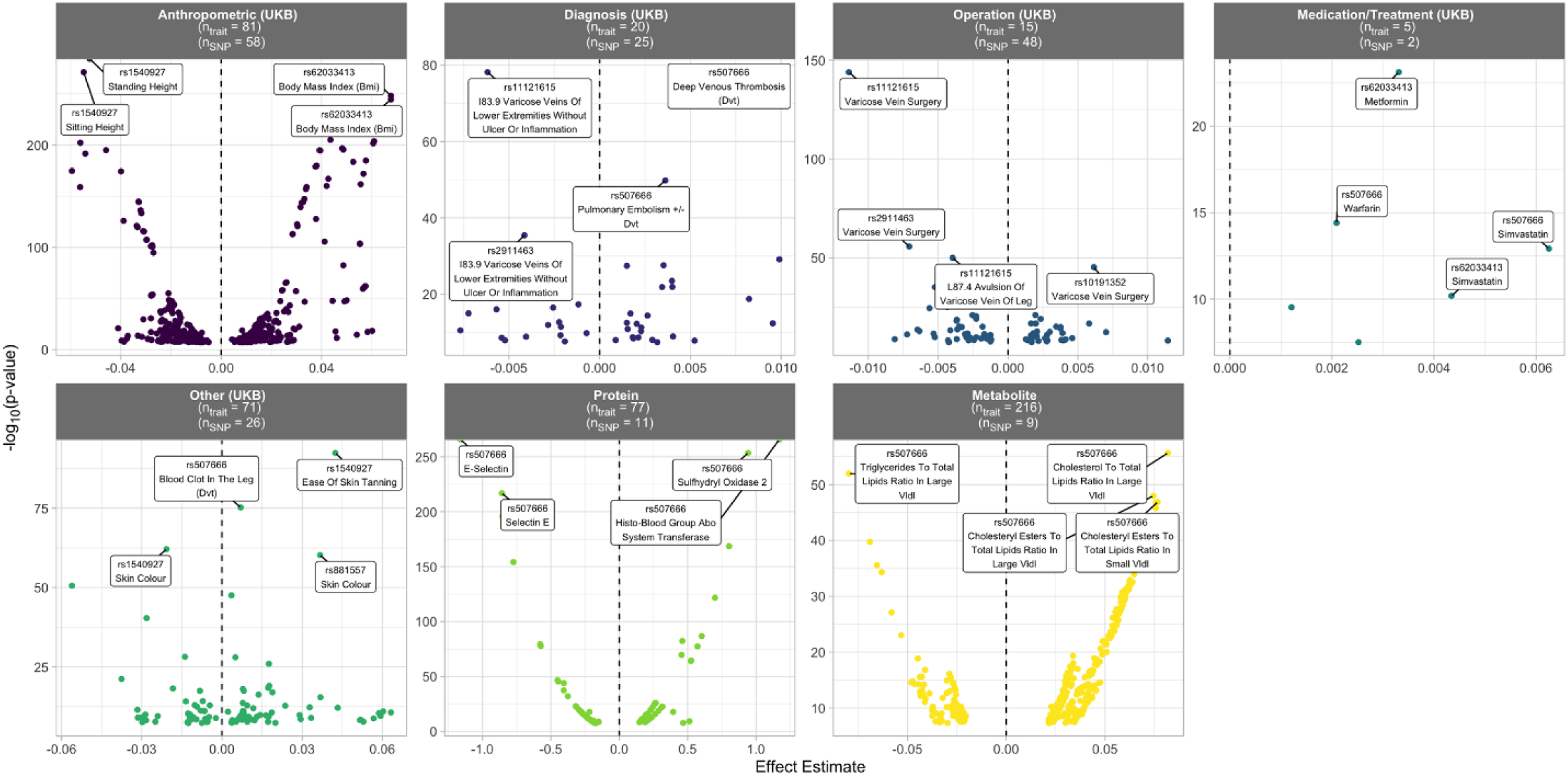
Phenome-Wide Associations of Varicose Veins Risk Variants. PheWAS was performed to identify associations between sentinel varicose veins-associated variants and traits with previously reported GWAS summary statistics available in the MRC IEU Open GWAS Project. Each points represents a genome-wide significant SNP-trait association (p < 5 × 10^−8^), with -log_10_(p-value) plotted as a function of the effect estimate. The top 4 associations by absolute z-score (z = effect/standard error) are labeled for within each category. The number of unique traits (n_trait_) sharing risk variants with varicose veins, and the number of unique varicose veins-associated SNPs (n_SNP_) associated with traits within each trait category are noted in the title of each facet.

Risk variants may be shared across traits based on linkage disequilibrium or shared causal genetic mechanisms.^24^ To prioritize traits that share causal genetic mechanisms with varicose veins, we performed Bayesian colocalization.^24^ We performed colocalization between each varicose veins risk locus (+/-500kb) and individual 1) UKB traits, 2) proteins, and 3) metabolites sharing genome-wide significant associations at that locus. We identified evidence of colocalization for 826 loci-trait pairs, across 70 distinct loci and 408 distinct traits. Anthropometric traits represented 46% (n = 72 unique traits at 37 loci) of UK Biobank traits sharing causal risk variants with varicose veins (n = 158 unique traits at 68 loci) (**FIGURE 3**). We also detected evidence of shared risk variants with circulating metabolites and proteins. These associations were concentrated at small number of pleiotropic loci (55 proteins at 4 loci, 195 metabolites at 6 loci) (**FIGURE 3**). At the *ABO* locus surrounding rs507666 (type A1 histo-blood group antigen),^25,26^ genes encoding 41 circulating proteins demonstrated evidence of colocalization with varicose veins, including several vascular proteins like tissue factor, E-selectin, ICAM1/2/4/5, PECAM1, P-selectin, and VEGFR2/3 among others.

**FIGURE 3:**
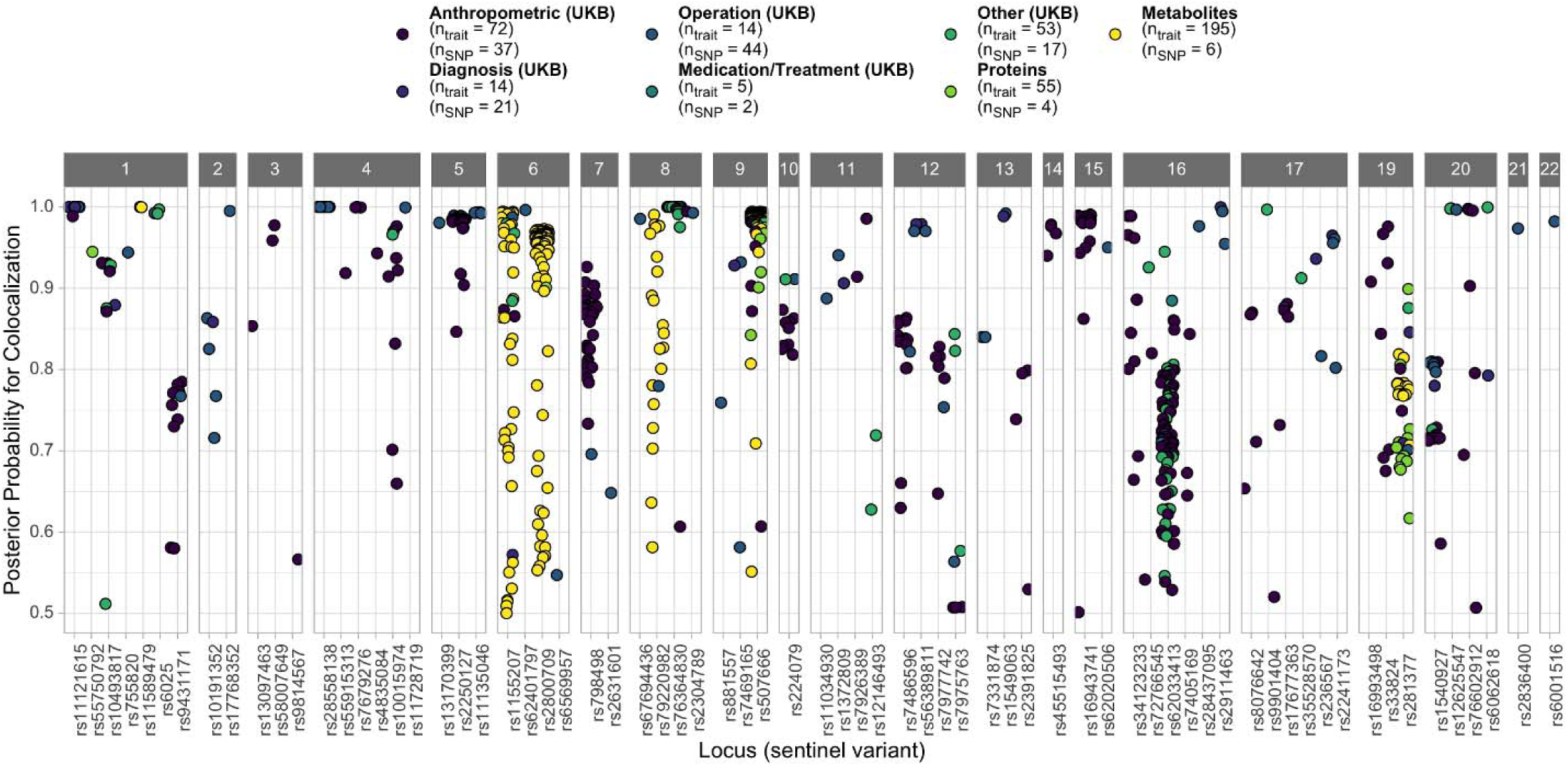
Bayesian Colocalization Reveals Shared Genetic Mechanisms. Bayesian colocalization was performed for the locus +/-500kb surrounding each significant pheWAS SNP-trait association. Each point represents a pairwise association between varicose veins and another trait, at a given locus (x-axis).

Next, multi-trait colocalization was performed, similarly testing for shared genetic mechanisms between varicose veins and clusters of 1) UKB traits, 2) proteins, and 3) metabolites (**SUPPLEMENTAL FIGURE 5**). We identified evidence of shared risk variants between varicose veins and UK Biobank traits at 34 loci (**SUPPLEMENTAL FIGURE 5**). We also found loci associated with varicose veins and clusters of circulating metabolites (3 loci) and circulating proteins (2 loci) (**SUPPLEMENTAL FIGURE 5**).

### Shared polygenetic architecture of vascular and anthropometric traits

While the shared genetic associations between varicose veins risk loci and anthropometric traits that were identified in pheWAS and colocalization analyses represent evidence of shared genetic architecture at the locus-level, varicose veins and anthropometric traits are largely polygenic traits.^21^ Therefore, we estimated genetic correlations between varicose veins, other vascular traits, and anthropometric traits to better understand shared polygenic patterns of genetic risk. A few notable findings emerged. Measures of height (eg. sitting height, standing height) were positively correlated with varicose veins and other vascular traits (eg. VTE and AAA), but negatively correlated with CAD (**FIGURE 4A**), consistent with prior reports.^16,27^ Measures of body composition (eg. body mass index, weight, fat mass, fat-free mass, body fat percentage) were largely positively correlated across all vascular traits. While systolic blood pressure was positively correlated with atherosclerotic traits like PAD, CAD, and stroke, we observed an inverse correlation with risk of varicose veins. Finally, markers of bone mineral density were uniquely correlated with PAD, but not other vascular traits.

**FIGURE 4:**
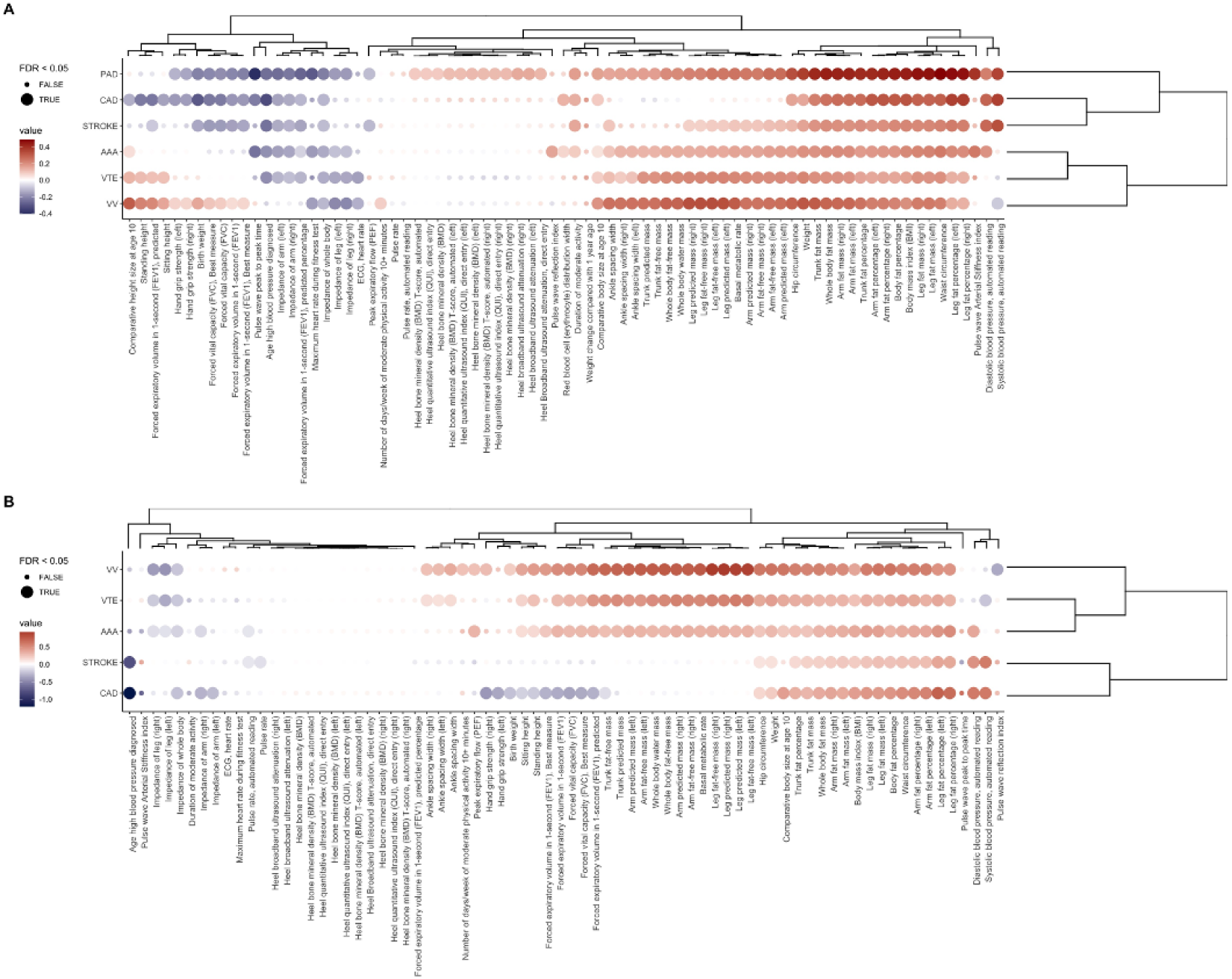
Shared Heritability of Anthropometric and Vascular Traits. Shared genetic architecture between varicose veins and anthropometric traits. A) Cross-trait LD-score regression was performed to evaluate the shared heritability between varicose veins, other vascular diseases, and anthropometric traits. B) Mendelian randomization was performed to evaluate whether anthropometric traits represent causal risk factors for varicose veins and other vascular diseases. Large circles represent significant findings (FDR < 0.05). Points are colored based on the direction and magnitude of genetic correlation (panel A), or direction of causal effect of each risk factor on each vascular trait (red = positive effect, blue = negative effect). Points are organized by hierarchical clustering of standardized effects (genetic correlation in panel A, z-score in panel B).

### Anthropometric traits as risk factors for varicose veins

To evaluate for putative casual effects of anthropometric traits on risk of varicose veins and other vascular disease, we performed inverse variance-weighted Mendelian randomization (**FIGURE 4B**). Measures of height (sitting height, standing height) and body composition (eg. body mass index, waist circumference, fat mass, fat-free mass, body water) were strongly associated with increased risk of varicose veins. Results were similar using the weighted-median MR method, which makes different assumptions about the presence of pleiotropy (**SUPPLEMENTAL TABLE 10**).

### Proteome-wide Mendelian randomization reveals circulating effectors of venous disease

We hypothesized that circulating proteins, which exist in close contact with the venous vasculature, may represent causal effectors of venous disease and thus novel treatment targets. To characterize the role of circulating proteins on risk of varicose veins, we performed proteome-wide Mendelian randomization. We selected previously established high-confidence genetic proxies of circulating proteins as instrumental variables within the Mendelian randomization framework.^28^ Of 714 circulating proteins with available *cis-*pQTL genetic proxies, 35 were significantly (FDR q < 0.05) associated with risk of varicose veins (**FIGURE 5; SUPPLEMENTAL TABLE 11**). These significant associations were enriched for proteins with molecular functions including extracellular matrix structure (FDR q = 1.2 × 10^−6^) and collagen binding (FDR q = 9.3 × 10^−3^); cellular components of the extracellular region (FDR q = 2 × 10^−9^); and biological processes including cell adhesion (FDR q = 9.1 × 10^−4^) and anatomical morphogenesis (FDR q = 1.2 × 10^−3^), among others (**SUPPLEMENTAL TABLES 12-14**). Examples of specific associations include adrenomedullin (*ADM*), a vasodilatory protein released by vascular endothelial cells in response to stress, which was associated with increased risk of varicose veins (OR 1.34, 95% CI 1.25 to 1.44, p = 1 × 10^−16^). Microfibrillar-associated protein 2 (*MFAP2*), an extracellular matrix protein which interacts with fibrillin (implicated in vascular diseases like Marfan syndrome), and associated with vascular phenotypes in zebrafish, was also associated with increased risk of varicose veins (OR 1.24, 95% CI 1.16 to 1.33, p = 2 × 10^−10^). We also identified proteins where higher circulating levels were associated with decreased risk of varicose veins, including Periostin (*POSTN*), a secreted extracellular matrix protein which binds integrins to support migration and adhesion of epithelial cells (OR 0.83, 95% CI 0.78 to 0.89, p = 4 × 10^−9^).

**FIGURE 5:**
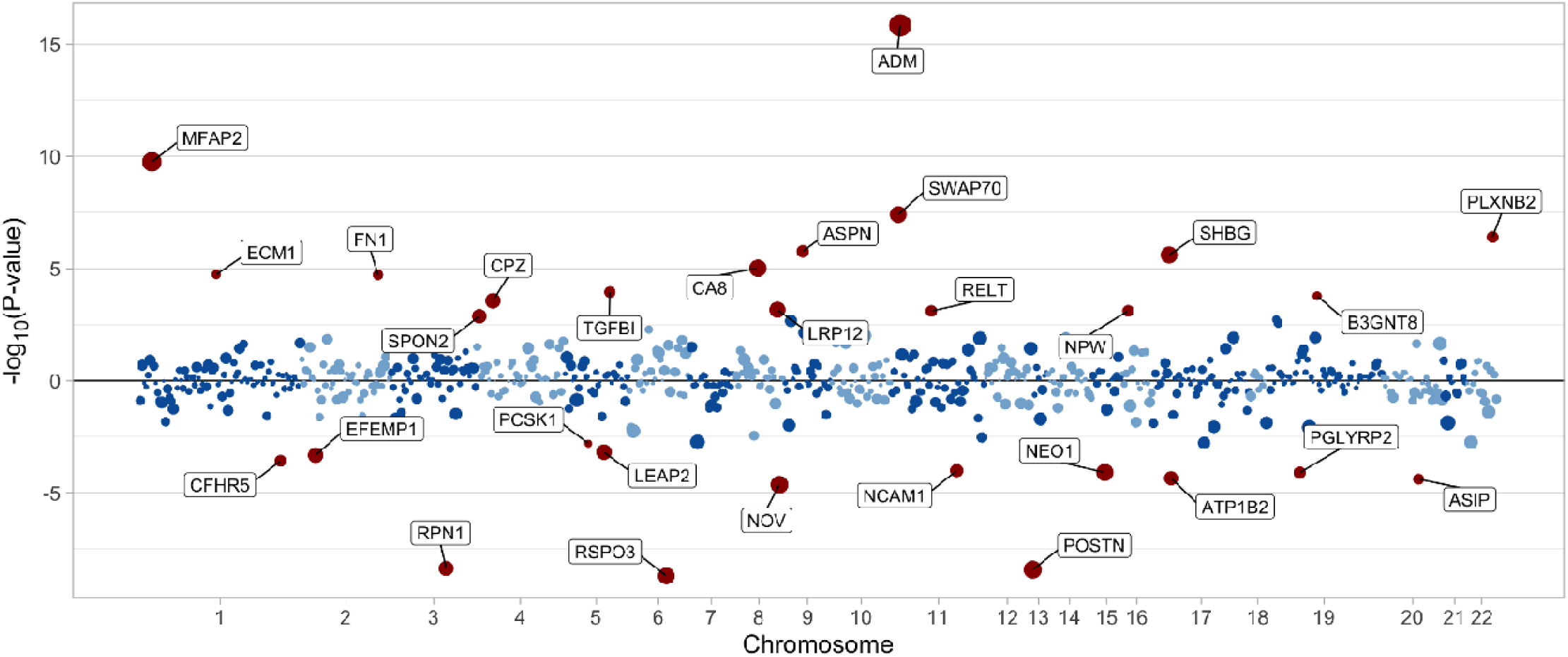
Proteome-wide Mendelian Randomization. Significant (FDR q < 0.05) associations between circulating proteins and varicose veins are highlighted in red and labeled with gene name. Size of points represents absolute effect size (larger points correspond to greater absolute effect size). Points above the origin correspond to proteins where higher circulating levels are associated with increased risk of varicose veins, while those below the origin correspond to proteins where higher circulating levels are associated with decreased risk of varicose veins.

### Drug-repurposing Mendelian randomization

We next evaluated whether drug targets of currently approved or clinical-stage therapeutics might be repurposed to modify risk of varicose veins. Two-sample Mendelian randomization was performed using previously defined genetic instruments proxying gene expression levels (*cis*-eQTLs) of drug targets in ChEMBL v.26.^29^ Of 983 actionable therapeutic targets with corresponding SNP effects for varicose veins, expression levels of 69 unique targets were significantly associated (FDR q < 0.05) with risk of varicose veins (**FIGURE 6; SUPPLEMENTAL TABLE 15**). Drug targets significantly associated with varicose veins included *PDE4C*, which encodes phosphodiesterase 4C. Although no medications are broadly available for treating varicose veins, pentoxifylline (a non-specific phosphodiesterase inhibitor) has been therapeutically applied for treatment of venous ulceration.^30^ Other drug targets with repurposing potential include *ITGB3* and *F2*, which encode integrin beta 3 and thrombin, respectively. Both targets implicate roles for thrombosis and coagulation, and can be therapeutically modulated using GPIIb/IIIa and direct-acting thrombin inhibitors, respectively.

**FIGURE 6:**
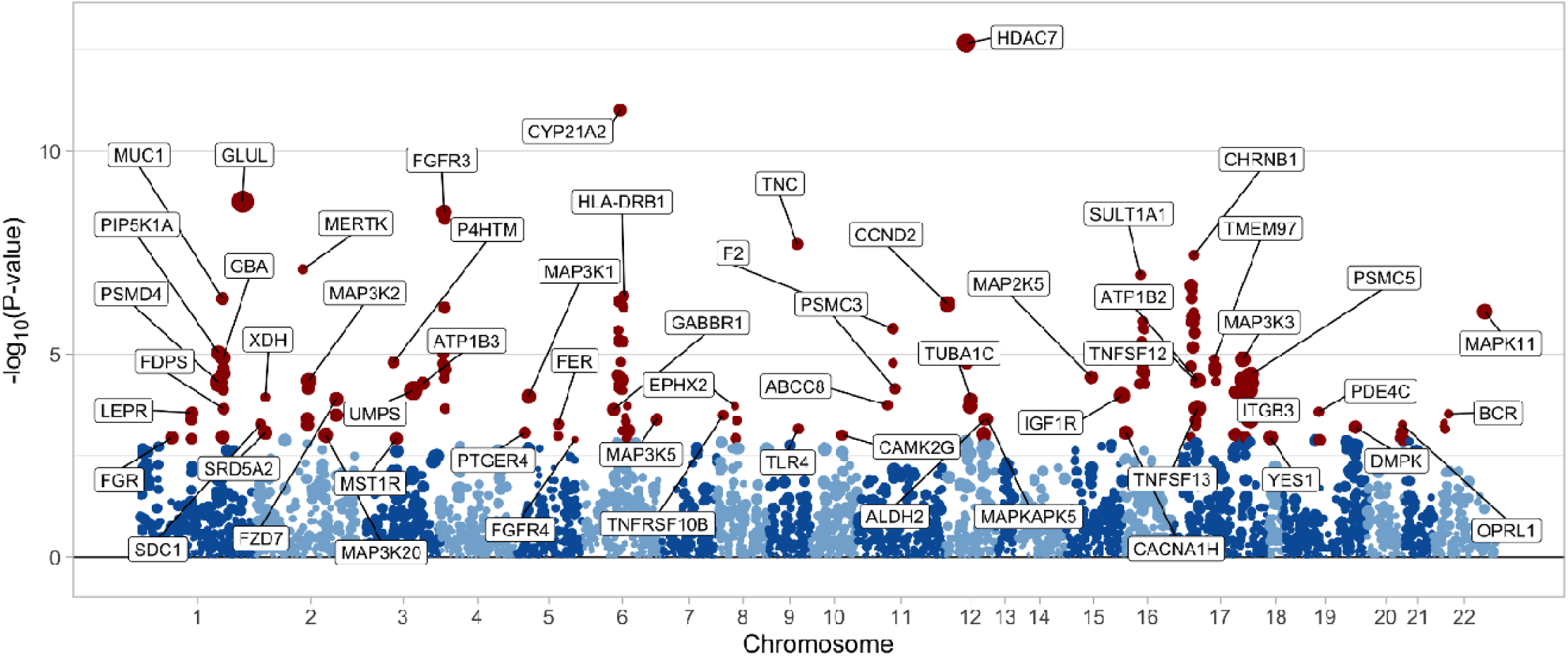
Drug-repurposing Mendelian Randomization. Significant (FDR q < 0.05) associations between gene expression levels of actionable drug targets and risk of varicose veins are highlighted in red and labeled with gene name. Size of points represents absolute effect size (larger points correspond to greater absolute effect size). Targets are positioned based on the chromosomal location of the protein-coding gene.

### Topological enrichment of varicose veins-associated biological pathways

Finally, we constructed an integrated mechanistic conceptual model of varicose vein pathophysiological processes (**FIGURE 7**). Genome-wide significant SNPs were mapped to genes using H-MAGMA (**SUPPLEMENTARY FIGURE 6A**) to integrate publicly available chromatin interaction data from human umbilical vein endothelial cells (GEO: GSE121520) with traditional proximity-based SNP-gene mapping.^31^ Genes were tested for standard enrichment of GO terms, as well as rank-based topological enrichment using random walk with restart (RWR) algorithms that traverse a multiplex network integrating 12 layers of omics data across relevant human tissues (**SUPPLEMENTARY FIGURE 6A**). We specifically considered gene enrichment in six GO biological processes associated with varicose veins pathophysiology: angiogenesis, blood pressure regulation, cytokine production, extracellular matrix (ECM) disassembly, leukocyte migration, and response to fluid shear stress. GWAS genes assigned by H-MAGMA were more topologically enriched in these six biological processes compared to random gene sets as determined by area under precision-recall curve (AUPRC; **SUPPLEMENTARY FIGURE 6B, 6C**). Furthermore, the GWAS genes were more topologically enriched for these six biological processes than nine GO biological processes not known to be associated with varicose vein pathophysiology (**SUPPLEMENTARY FIGURE 6C**).

**FIGURE 7:**
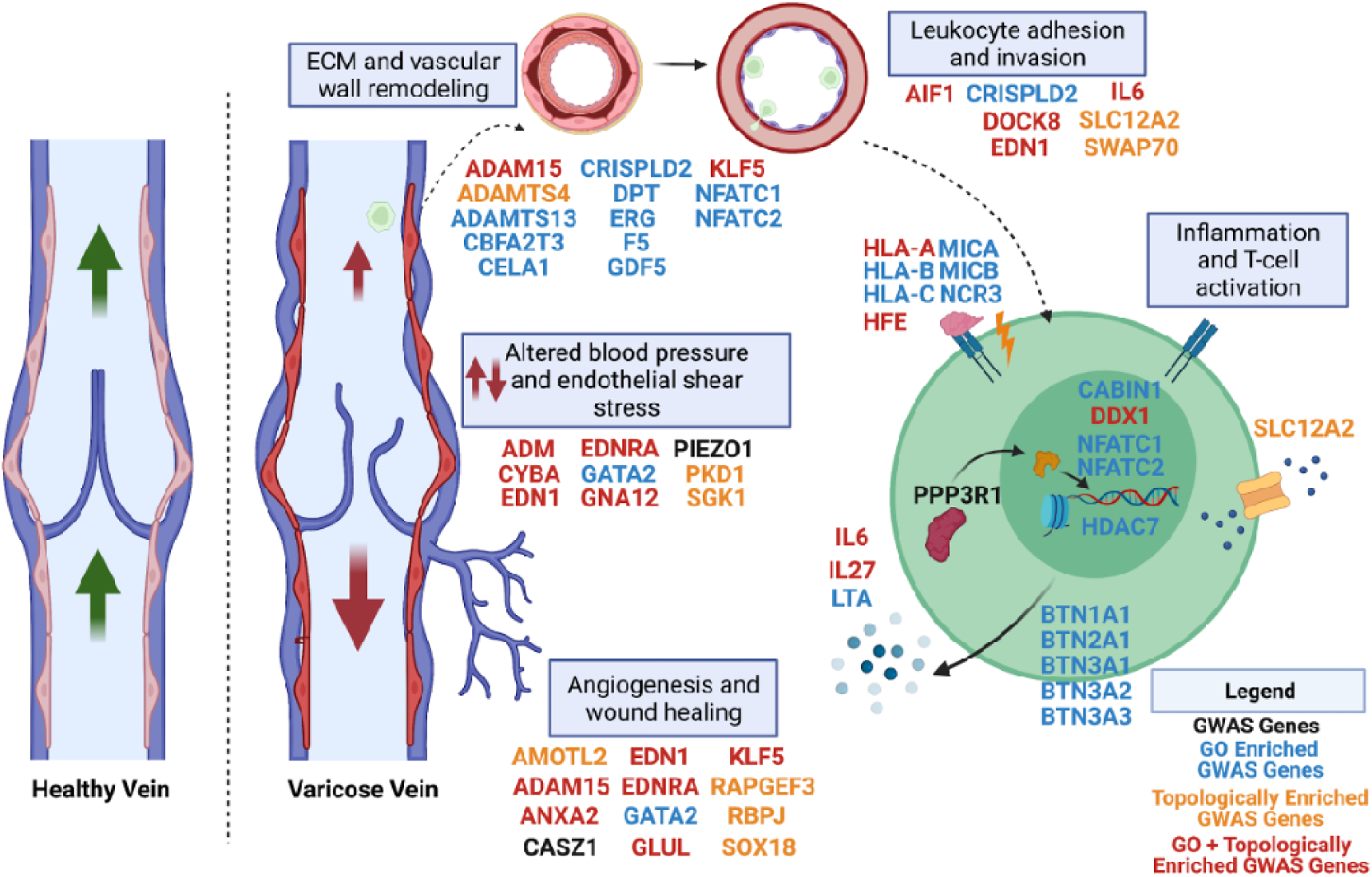
Systems biology integration of genome-wide significant variants contributing to varicose vein pathophysiology. Known varicose vein pathophysiological processes are implicated by GWAS genes as determined by H-MAGMA (black). GWAS genes also overlapped with traditional GO enrichment (blue), topological enrichment of GO terms (orange), or both GO enrichment and topological enrichment (red). Multiple genes were implicated in angiogenesis and dysregulated blood pressure including GATA2, endothelin-1 (EDN1) and the endothelin A receptor (EDNRA). Vascular wall and extracellular matrix (ECM) remodeling genes were also enriched including ADAM15, ADAMTS4, and ADAMTS13. Variants in leukocyte migration genes were identified, including AIF1 and DOCK8. Finally, pro-inflammatory and T-cell activation genes were also enriched (CABIN1, IL6, IL27, LTA, NFATC1, NFATC3, and PPP3R1) as well as major histocompatibility complex class 1 (MHC-I) and butyrophilin genes.

As seen in the mechanistic conceptual model depicted in **FIGURE 7**, using gene set and topological enrichment we identified GWAS genes related to altered blood pressure and endothelial shear stress (9 genes), angiogenesis and wound healing (12 genes), ECM and vascular wall remodeling (13 genes), leukocyte adhesion and invasion (7 genes), and inflammation and T-cell activation (23 genes). We highlight several examples of enriched gene sets which recapitulate molecular mechanisms of varicose veins. For example, endothelin-1 (*EDN1*), the endothelin-A receptor (*EDNRA*), and the transcription factor *GATA2*, are known to contribute to both angiogenesis and hypertension in varicose vein patients through a vascular endothelial growth factor (VEGF)-mediated mechanism that is sensed by *PIEZO1*.^32–35^ The transcription factor *KLF5* similarly promotes angiogenesis by its transcriptional activation of *VEGFA* (one of the genes encoding VEGF) and increases vascular wall remodeling by causing transcription of the matrix metalloprotease genes *MMP2* and *MMP3*.^36^ Additional matrix metalloproteases (*ADAM15, ADAMTS4, ADAMTS13*) associated with increased endothelial permeability in venous disease were also implicated.^37–40^ Immune-related genes included *DOCK8* (dendritic cell migration), *AIF1* (dendritic cell differentiation and macrophage migration), and several genes implicated in T-cell activation and inflammatory processes including *PPP3R1* (calcineurin subunit), *CABIN1* (calcineurin binding protein), two *NFAT* subunits (*NFATC1* and *NFATC2*), MHC class I genes (*HLA-A, HLA-B, HLA-C, MICA*, and *MICB*) that lead to antigen presentation to T-cells, and several butyrophilin genes (*BTN1A1, BTN2A1, BTN3A1, BTN3A2, BTN3A3*) which modulate T-cell cytokine release.^41–45^ Finally, the pro-inflammatory cytokines *IL6* and *LTA* were identified in tandem with *IL27*, a cytokine that induces T-cell differentiation and has been implicated in cardiovascular disease more broadly.^46^ These topological enrichment findings demonstrate that genetic variants present in varicose vein patients influence the major pathophysiologic processes that arise in the course of disease.

## DISCUSSION

This GWAS meta-analysis represents the largest multi-ancestry analysis of varicose veins to date. Overall, we identified 139 risk loci, establishing novel credible risk variants. We characterized the shared genetic architecture of varicose veins with other traits at the locus-and polygenic levels. Mendelian randomization defined putative casual clinical risk factors and identified circulating proteins and drug targets that may represent causal treatment targets. Finally, our integrative topological enrichment analysis systematically linked human genetic data with major biological processes implicated in varicose veins.

First, our findings build on prior studies demonstrating the heritability of varicose veins and extend these findings using larger cohorts of diverse genetic ancestry. Our identification of 139 varicose veins risk loci represents a 3.8-fold improvement in genomic discovery compared with prior reports, likely a result of increased sample size and cohort diversity. These findings are broadly consistent with similarly large and diverse genome-wide analyses of complex traits.^47^ Further, we find evidence of genetic overlap between both venous and arterial disease. This finding is supported at the polygenic level by significant genetic correlation between varicose veins and both arterial (peripheral artery disease and abdominal aortic aneurysm), and venous disease (venous thromboembolism). We demonstrate additional overlap at the locus-level. For instance, F5 p.Arg534Gln (factor V Leiden), a commonly-recognized risk factor for venous thromboembolism, is now identified as a novel risk locus for varicose veins. Recent evidence further suggests this risk variant contributes to risk of peripheral artery disease, blurring the traditional differences in arterial and venous risk factors.^48^

Second, our findings implicate pathways relevant to both primary and secondary etiologies of varicose veins. Clinically, varicose veins are classified based on the CEAP classification scheme, where varicose veins occur based on congenital, primary, secondary, or no identifiable venous etiologies.^9^ Although we did not detect substantial overlap with Mendelian diseases associated with varicose veins, we did find genetic architecture relevant to both primary and secondary etiologies. Focusing on genome-wide significant loci, both pathway enrichment and colocalization analyses implicated vascular pathways and proteins relevant to primary varicose veins. For example, our prioritization of vascular smooth muscle cell pathways is consistent with *in vitro* evidence finding dysregulated smooth muscle cell phenotypes in varicose veins in comparison to healthy control vessels.^49^ Similarly, MAGMA tissue and pathway enrichment at the polygenic level highlighted vascular tissues and pathways, while Mendelian randomization identified causal vascular proteins and drug targets. At both the locus and polygenic levels we also identified genetic overlap with secondary causes of varicose veins (Eg. the *FTO* locus and genetic correlation with anthropometric markers like obesity; the F5 locus and genetic correlation with venous thromboembolism). While these findings support biological pathways relevant to the current varicose veins classification scheme, future study will be required to determine whether increased polygenic risk specifically within vascular pathways is necessary/sufficient for disease, and whether gene-environment interactions (particularly obesity) alter disease risk.

Third, our findings highlight the pleiotropic effects of obesity and related anthropometric traits. Our genetic correlation and Mendelian randomization analyses demonstrate that vascular and anthropometric traits are closely linked, and that body composition adversely influences vascular disease risk more broadly. While some markers of obesity and body composition have been previously linked to varicose veins,^16,18^ we extend those findings to demonstrate shared pattens of risk across common vascular diseases. Although varicose veins (in contrast to diseases like coronary/peripheral artery disease) are not likely to be the driving force behind public health efforts to curb obesity, our findings suggest that targeting obesity and related markers of body composition would be expected to generally improve the vascular risk profile, reducing the burden of varicose veins as a beneficial consequence.

Fourth, we identify novel pathways relevant to the pathogenesis of varicose veins. For example, we identify a novel risk locus (rs55750792) near microfibrillar associated protein 2 (MFAP2; also known as MAGP1). Our protein Mendelian randomization analysis identified a strong association between circulating levels MFAP2, and increased risk of varicose veins, a finding reinforced by significant colocalization between varicose veins and circulating MFAP2 (posterior probability 0.95). MFAP2 is an extracellular matrix protein with a carboxy-terminal fibrillin and tropoelastin-binding domains and an amino-terminal domain capable of binding active transforming growth factor beta (TGF-β).^50–52^ Several plausible mechanisms link MFAP2 to risk of varicose veins. At the extracellular matrix, disorganized fibrillin distribution and increased TGF expression have been observed within varicose veins in comparison with health control vessels.^53^ Consistent with these findings, we observed a novel risk variant (rs12123737) at the TGFBR3 locus, and find that genetically-proxied circulating levels of TGFBI (TGF-β induced protein) were also associated with increased risk of varicose veins. Prior observations found significantly increased TGFBI levels in varicose veins in comparison with normal control vessels.^54^ MFAP2 also binds NOTCH1, and crosstalk between notch, TGF, and vascular endothelial growth factor pathways play key roles in vascular morphogenesis. We identify a novel varicose veins risk locus (rs507666), which also represents a pleiotropic *trans*-pQTL for members of the vascular endothelial growth factor receptor family (VEGFR2 and VEGFR3). Further, rs72775784 at the NOTCH1 locus represents another novel risk variant for varicose veins. In addition to the functions of MFAP2 in the extracellular matrix and vascular morphogenesis which may lead to primary varicose veins, recent evidence suggests MAFP2 binds several hemostatic factors including fibronectin, fibrinogen, and von Willebrand factor, contributing to hemostasis and thrombosis, suggesting an additional mechanism for MFAP2 relevant to secondary causes of varicose veins.^55^

Fifth, we identify drug-repurposing opportunities, providing human genetic evidence to motivate additional mechanistic studies and clinical trials. For example, our prioritization of *PDE4C* provides human genetic evidence to study the role of pentoxifylline for the treatment of varicose veins, while prioritization of obesity (eg. *LEPR*), thrombosis (eg. *ITGB3*), and coagulation (eg. *F2*) targets reinforce the role of these pathways for secondary forms of varicose veins. Future mechanistic evaluation of these targets for varicose veins is warranted, however the genetic evidence for these pathways increases the likelihood of successful clinical translation.^56,57^

Finally, our systems biology-inspired enrichment analysis expands the number of loci linked to biological processes known to be disrupted in varicose vein pathology. By leveraging tissue/cell-type specific epigenetic date, we refine SNP-to-gene assignment compared to methods applied in previous varicose vein GWAS.^16,18^ By combining standard GO enrichment with a novel RWR approach, we demonstrate that genes from the present GWAS are enriched for functional connectivity to biological pathways important to varicose veins. This novel gene set enrichment analysis differs from standard gene set enrichment in that it does not rely entirely on the intersection of GWAS genes with genes in a GO term. Instead, enrichment is determined by demonstrating that both GWAS genes and genes pertaining to a GO term exhibit similar network topological connectivity across a multiplex network integrating 12 layers of multi-omics data using ranks determined by RWR, indicating that they are involved in similar biological processes. Thus, by using our network-based approach, here we link genetic variants from GWAS into a systems-level integration of biological pathways involved in varicose vein pathophysiology.

### Limitations

Our findings should be interpreted within the context of several limitations. First, we sought to assemble the broadest and most diverse set of individuals with varicose veins to improve power for genetic discovery, recognizing that concurrent formal replication in a similarly diverse and well-powered cohort would be unlikely. However, we apply complementary colocalization, genetic correlation, Mendelian randomization, and systems biology methods to prioritize loci and pathways for further study.^58^ Second, our definition of varicose veins was based on prevalent diagnosis codes ascertained from the electronic health record. Although several varicose veins classifications/subtypes have been described based on different pathophysiologic mechanisms, identifying varicose veins subtypes at scale using the electronic health record remains limited. While we identify pathways relevant to both primary and secondary subtypes of varicose veins, interactions between these pathways and their relevance across individual subtypes requires further study. Further, the VA Million Program cohort is >90% male, and we do not explore sex-specific differences. While Mendelian randomization has successfully predicted the therapeutic effects of drug targets, the method formally proxies lifelong genetic exposure rather than short-term pharmacologic interventions. The therapeutic effect sizes predicted in our analyses may therefore differ in comparison to pharmacologic treatment.

### Conclusion

In the largest and most diverse genome wide association study of varicose veins to-date, we identify novel genetic risk loci for varicose veins, highlight shared risk factors with other vascular diseases, connect genetic variants to biological pathways with multi-omic network analysis, and identify novel therapeutic targets and drug repurposing opportunities.

## METHODS

### Study Populations

Individuals of diverse ancestry with and without varicose veins were identified using electronic health record diagnosis codes linked to genetic data from the VA Million Veteran Program,^20^ UK Biobank,^59^ eMERGE (https://emerge-network.org), FinnGen (https://finngen.gitbook.io/documentation/), and BioBank Japan (http://jenger.riken.jp/en/). The VA Million Veteran program recruits adults age >18 from Veterans Affairs Medical Centers across the USA. From MVP, we evaluated 18,977 cases (15,240 European-ancestry; 2511 African-ancestry; 1,226 Hispanic-ancestry) and 561,203 controls (409,037 European-ancestry; 106,998 African-ancestry; 45,168 Hispanic-ancestry) from Release 4. From UK Biobank, eMERGE, FinnGen, and BioBank Japan, we evaluated an additional 30,788 cases and 773,098 controls. Details of cohorts and phenotype definitions are available in the **Supplemental Methods**. All studies received ethical and protocol approval from the relevant institutional review boards, and all participants provided informed consent. The VA Million Veteran Program was approved by the VA Central Institutional Review Board

### Genome Wide Association Study Meta-Analysis

Details of study-specific genotyping and quality control are available in the **Supplemental Methods**. Briefly, ancestry-specific genome wide association studies were performed within each cohort, including covariates for age, sex, and population structure. MVP analyses were performed using logistic regression in plink2a,^60,61^ eMERGE analyses were performed using regenie,^62^ and publicly-available GWAS from UK Biobank, FinnGen and BioBank Japan were performed using SAIGE,^63^ which are computationally-efficient methods for performing GWAS within biobank-scale populations. GWASinspector^64^ was used to perform study-level quality control (using default settings), and to harmonize effect alleles across studies. Fixed-effects inverse variance-weighted GWAS meta-analysis was performed using METAL,^65^ first withi- and then across ancestries to generate a multi-ancestry meta-analysis. Independent variants and loci associated with varicose veins were identified using FUMA v1.3.6d (https://fuma.ctglab.nl/),^66^ using a genome wide-significance threshold of p < 5 × 10^−8^, and default parameters for pruning independent variants/loci (merge-distance 250kb; linkage disequilibrium r^2^=0.6, r_2_^2^ = 0.1). All genomic positions are reported using coordinates from the GRCh37 build of the human genome.

### Fine-mapping

Multi-ancestry fine-mapping was performed using MR-MEGA.^23^ This statistical method performs meta-regression, using principal components to account for heterogeneity in allelic effects that is correlated with ancestry. We considered study-level associations at each genome-wide significant locus, using the default settings to account for 4 genetic principal components. At each genome-wide significant locus (+/-500kb of each lead variant) we derived 99% credible sets by ranking variants by Bayes’ factor and including variants until the cumulative posterior probability reached 0.99.

### Tissue and Gene-set Enrichment

Tissue and gene-set enrichment were evaluated using MAGMA v1.08, via the FUMA web application.^66,67^ MAGMA aggregates SNP-level associations into gene-level test statistics. For tissue enrichment analysis, MAGMA performs gene property analysis, accounting for average tissue expression across 54 tissues (identified using GTEx v8).^68^ For gene-set enrichment, 15,485 gene sets (“Curated gene sets” and “GO terms” from MSigDB v7.0)^69^ were tested for enrichment with varicose veins risk loci.

### Phenome-wide association study (pheWAS)

Lead variants at each of the 139 loci significantly associated with varicose veins were queried using the UK Medical Research Council Integrative Epidemiology Unit OpenGWAS project (https://gwas.mrcieu.ac.uk/).^70,71^ We retrieved associations between these 139 varicose veins-associated variants and traits for which prior genome-wide association studies had been performed, focusing on UK Biobank diagnoses/operations/medications/surveys/measurements (n = 2514; from the “ukb-b” dataset), circulating proteins (n = 4489; from the “prot-” datasets), and GWAS of circulating metabolites (n = 974; from the “met-” datasets). Given some overlap of proteins and metabolites across individual datasets, we considered the conventional genome-wide association study significance threshold of p < 5 × 10^−8^, which is similar to p-value threshold of 0.05/[139*(2514+4489+974)] when using the Bonferroni adjustment for multiple testing.

### Colocalization

Bayesian colocalization was performed using the default parameters of HyPrColoc.^24^ HyPrColoc enables colocalization between arbitrary-sized groups of traits, remaining computationally efficient even when evaluating hundreds of traits simultaneously. We performed colocalization between each varicose veins risk locus (+/-500kb) and traits that shared risk loci (identified using our pheWAS). Colocalization was first performed pairwise, individually testing for colocalization between varicose veins and each trait sharing a risk locus. Next, multi-trait colocalization was performed, simultaneously evaluating for colocalization between varicose veins and all traits sharing a given risk locus. The default posterior probability threshold P_R_P_A_ > 0.49 was used to identify significant colocalization.

### Genetic Correlation

Genetic correlations were estimated using cross-trait LD-score regression, using LDSC v1.0.1 (https://github.com/bulik/ldsc).^21,72^ When estimating correlations with vascular traits, summary statistics were obtained from large genome wide association studies of coronary artery disease, peripheral artery disease, ischemic stroke, venous thromboembolism, and abdominal aortic aneurysm.^48,73–76^ When estimating correlations with anthropometric traits, summary statistics were obtained from publicly-available genome-wide association studies of anthropometric traits measured among UK Biobank participants, identified from the “ukb-b” dataset of the MRC-IEU OpenGWAS project (https://gwas.mrcieu.ac.uk/).^70,71^

### Anthropometric Mendelian Randomization

Genetic instruments for anthropometric traits were constructed from independent (r^2^<0.001, distance = 10,000kb) genetic variants associated with each anthropometric trait at genome-wide significance (p < 5 × 10^−8^), from publicly available GWAS (described in Genetic Correlation section above). Corresponding SNP effects in each vascular trait (varicose veins, coronary artery disease, peripheral artery disease, venous thromboembolism, and abdominal aortic aneurysm) were identified, and harmonized to ensure concordant effect alleles. Inverse variance-weighted two-sample Mendelian randomization was then performed using the *TwoSampleMR* package in R.^70^ As a sensitivity analysis we applied the weighted-median method, which makes different assumptions about the presence of pleiotropy and invalid instruments.^77^ To account for multiple testing, the false discovery rate was controlled at 5%.

### pQTL Mendelian Randomization

Details of genetic instrument selection and validation have been previously reported.^28^ Briefly, we considered high-confidence (tier 1) genetic instruments which were identified based on independent (r^2^ < 0.001) cis-pQTLs which were associated with circulating protein levels at a GWAS p-value threshold ≤⍰5⍰× ⍰10^−8^, and passed both specificity and consistency tests.^28^ Of genetic instruments for 738 circulating proteins, corresponding varicose veins outcome data was available for 714 (96.7%). For each of the 714 genetic instruments (each representing a single locus/protein), Mendelian randomization was performed using the Wald ratio method.^78^ To account for multiple testing, the false discovery rate was controlled at 5%.

### Drug-Repurposing Mendelian Randomization

Details of genetic instrument selection have been previously reported.^29^ Briefly, genes encoding the targets of approved or clinical-stage therapeutics were identified using ChEMBL v.26.^79^ Conditionally-independent *cis*-eQTLs for each gene was then identified from GTEx v8.^68^ When multiple instruments from different GTEx v8 tissues were present, each tissue was tested independently. Two sample Mendelian Randomization was then performed to estimate the effect of transcript levels of each drug target on varicose veins. Inverse-variance weighting was used to combine effects when multiple genetic instruments for a transcript were available. To account for multiple testing, the false discovery rate was controlled at 5%.

### Hi-C data processing and H-MAGMA SNP-to-gene assignment

Hi-C data from control human umbilical vein endothelial cells (HUVECs) were downloaded from publicly available data (GEO: GSE121520)^80^ aligned to reference genome hg19/GRCh37. Significant chromatin contacts were determined at 5kb resolution using Fit-Hi-C using 2 spline passes with lower bound threshold of 10kb, upper bound threshold of 2Mb, and significant contacts defined as *p* < 0.01.^81,82^ We used this information to generate an annotation file for H-MAGMA for HUVEC-specific SNP-to-gene assignment.^31^ 8,123 SNPs exceeding genome-wide significance (p < 5 × 10^−8^) were included for H-MAGMA gene assignment, resulting in 638 unique genes identified (Supplementary Table 16).

### Multiplex network generation

A multiplex network (a homogenous multilayer network where individual layer topologies are retained) containing gene nodes was created from 12 layers of omics data using the RandomWalkRestartMH R package.^83^ Protein-protein interactions were merged into a single network layer from HumanNetv2 and STRING (v11) databases.^84,85^ Co-citation, co-essentiality, co-expression, pathway database, gene neighborhood, interlog, and phylogenetic network layers from HumanNetv2 were also included as individual layers. Transcription factor - target networks were downloaded from hTFtarget^86^ on 2/20/2021 and merged into a single network layer derived from the following 15 HUVEC ChIP-seq GEO datasets: BRD4, RELA: GSE53998;^87^ CTCF: GSE29611;^88^ ELK3: GSE60156;^89^ EP300, ETS1: GSE41166;^89^ EWSR1, FLI1: GSE31838;^90^ FOS, GATA2, JUN, MAX: GSE31477;^88^ MYC: GSE33213; PGR: GSE43786;^91^ PPARD/PPARG: GSE50144.^92^ Three tissue type-specific predictive gene expression networks were generated using the explainable-AI algorithm iterative random forest leave one-out prediction (iRF-LOOP)^93,94^ from the following GTEx v8 RNA-seq data sets downloaded from the GTEx Portal on 10/13/2020: tibial artery, whole blood/EBV-transformed lymphocytes, and liver.^95^

### Network-based topological functional enrichment

The set of significantly associated GWAS genes from H-MAGMA was tested for enrichment for genes belonging to six key Gene Ontology (GO) annotations by evaluating their connectivity in the multiplex network: GO:0001525: angiogenesis; GO: 0002367: cytokine production involved in immune response; GO:0008217: regulation of blood pressure; GO:0022617: extracellular matrix disassembly; GO:0034405: response to fluid shear stress; GO:0050900: leukocyte migration. The entire GWAS geneset was used as seed genes for the Random Walk with Restart (RWR) algorithm, producing a ranking of all other genes in the multiplex according to their joint topological connectivity to the GWAS genes. Area Under Precision Recall Curve (AUPRC) was calculated and used to score the ability for the random walker to effectively find each target geneset when starting from the GWAS genes. The same was done for 20 random genesets of the same size as each target gene set to determine if the score was better than random. Nine non-target, brain-related GO biological process gene sets were also scored for comparison purposes: GO:0001963: synaptic transmission, dopaminergic; GO:0007411: axon guidance; GO:0021766: hippocampus development; GO:0021854: hypothalamus development; GO:0035249: synaptic transmission, glutamatergic; GO:0042165: neurotransmitter binding; GO:0051932: synaptic transmission, GABAergic; GO:0098664: G protein-coupled serotonin receptor signaling pathway; GO:1903859: regulation of dendrite extension. GWAS genes were also tested for standard GO enrichment using ToppGene (https://toppgene.cchmc.org/).^96^ GO-enriched and topologically enriched gene lists from H-MAGMA genes are included in **Supplementary Tables 17-18**.

## Data Availability

GWAS meta-analysis summary statistics will be deposited in dbGaP upon final publication. GWAS summary statistics from FinnGen, BioBank Japan, and UK Biobank are publicly available from https://r4.finngen.fi/, https://pheweb.jp/, and https://pan.ukbb.broadinstitute.org.

## ACKNOWLEDGEMENTS

The authors thank the participants of the VA Million Veteran Program, UK Biobank, eMERGE, FinnGen, and BioBank Japan studies.

## FUNDING

MGL is supported by the Institute for Translational Medicine and Therapeutics of the Perelman School of Medicine at the University of Pennsylvania and the NIH/NHLBI National Research Service Award postdoctoral fellowship (T32HL007843). SMD is supported by the US Department of Veterans Affairs award #IK2-CX001780. This publication does not represent the views of the Department of Veterans Affairs or the United States Government. SMD receives research support from RenalytixAI and personal consulting fees from Calico Labs, both outside the scope of the current manuscript. DAJ, KAS, DK, MRG, ML, AMC, and JR are supported by funding from the Million Veteran Program Computational Health Analytics for Medical Precision to Improve Outcomes Now (CHAMPION) initiative and NIH grants DA051908 and DA051913. eMERGE is supported by U01HG8657 (University of Washington); U01HG8685 (Brigham and Women’s Hospital); U01HG8672 (Vanderbilt University Medical Center); U01HG8666 (Cincinnati Children’s Hospital Medical Center); U01HG6379 (Mayo Clinic); U01HG8679 (Geisinger Clinic); U01HG8680 (Columbia University Health Sciences); U01HG8684 (Children’s Hospital of Philadelphia); U01HG8673 (Northwestern University); U01HG8701 (Vanderbilt University Medical Center serving as the Coordinating Center); U01HG8676 (Partners Healthcare/Broad Institute); and U01HG8664 (Baylor College of Medicine). This research used resources of the Oak Ridge Leadership Computing Facility, which is a DOE Office of Science User Facility supported under Contract DE-AC05-00OR22725. This manuscript has been co-authored by UT-Battelle, LLC under contract no. DE-AC05-00OR22725 with the U.S. Department of Energy. The United States Government retains and the publisher, by accepting the article for publication, acknowledges that the United States Government retains a nonexclusive, paid-up, irrevocable, world-wide license to publish or reproduce the published form of this manuscript, or allow others to do so, for United States Government purposes. The Department of Energy will provide public access to these results of federally sponsored research in accordance with the DOE Public Access Plan (http://energy.gov/downloads/doe-public-access-plan, last accessed September 16, 2020).

## Notes

### Author Declarations

The VA Million Veteran Program was approved by the VA Central Institutional Review Board

