## Supplemental Methods and Figures for "Multi-ancestry Genome-wide Association Study of Varicose Veins Reveals Polygenic Architecture, Genetic Overlap with Arterial and Venous Disease, and Novel Therapeutic Opportunities"

#### Study Populations and Phenotyping

VA Million Veteran Program: The VA Million Veteran Program is a mega-biobank which has recruited >825,000 adult (age >18 years) participants from Veterans Affairs Medical Centers across the United States.[1] Participants consent to linkage of electronic health record and genomic data, including diagnosis and procedure codes, laboratory measurements, radiology images/reports, and others. Participants also complete a baseline assessment and survey. Individuals with varicose veins were identified using electronic health record diagnosis codes (ICD9/10) mapped to pheCodes.[2] Individuals with 2 or more instances of pheCode 454.1 were classified as varicose veins cases, while individuals with 0 instances were considered controls. Individuals with 1 instance were excluded. Individuals of diverse genetic ancestry, determined using HARE, were included.[3] MVP received ethical and study protocol approval by the VA Central Institutional Review Board and informed consent was obtained from all participants.

UK Biobank: The UK Biobank is a prospective cohort study which enrolled >500,000 participants age 60-69 years from across the United Kingdom.[4] Prospective data collected on participants includes physical measures, surveys, laboratory testing, and imaging, with additional linkage to national health records and registries including diagnoses, procedures, medications, and measurements, among others. Individuals with varicose veins were identified using pheCode 454.1. The UK Biobank has ethical approval from the North West Multi-centre Research Ethics Committee (MREC; 11/NW/0382), the Patient Information Advisory Group (PIAG) and National Information Governance Board for Health and Social Care (NIGB) in the UK, and the Community Health Index Advisory Group (CHIAG) in Scotland.

eMERGE: The Electronic Medical Records and Genomics network (https://emerge-network.org) was established in 2007 and represents a network of biorepositories linked to electronic health records for the purposes of studying and implementing genomic medicine. Individuals with varicose veins were identified using pheCode 454.1. Participants in eMERGE provide informed consent, and the network has received ethical approvals at participating institutions.

BioBank Japan: BioBank Japan is a nationwide hospital-based biobank which enrolled ~200,000 participants from 66 hospitals affiliated with 12 medical centers across Japan between 2003-2008. Clinical information including surveys, health records, and serum samples were obtained from patients annually until 2013. Individuals with varicose veins were identified using the ICD10 code I83, corresponding to pheCode 454.1. The BioBank Japan Project was approved by the research ethics committees at the Institute of Medical Science, the University of Tokyo, the RIKEN Yokohama Institute, and cooperating hospitals; participants gave written informed consent.

FinnGen: FinnGen is a public-private partnership which aims to collect genome and health data on 500,000 Finnish biobank participants. The study consists of ~200,000 legacy samples primarily collected by the National Institute for Health and Welfare, and an additional ~300,000 samples to be prospectively collected from hospital biobanks. Participating individuals consent to linkage of genome-wide genotyping with nationwide registers of longitudinal health data. Individuals with varicose veins were identified using electronic health record diagnosis codes ICD9 454 or ICD10 I83, which correspond to pheCode 454.1. FinnGen participants provided informed consent for biobank research, and the Coordinating Ethics Committee of the Hospital District of Helsinki and Uusimaa (HUS) approved the FinnGen Study protocol No. HUS/990/2017.

#### Genotyping and Quality Control

VA Million Veteran Program: DNA was extracted from whole blood and genotyped using a customized Affymetrix Axiom biobank array. Duplicate samples and those with high heterozygosity, missing genotypes, sex discordance, Hardy-Weinberg Equilibrium $p < 1 \times 10^{-6}$, and relatedness (as measured by KING[5]) were excluded. Phasing was performed using SHAPEIT4,[6] and minimac4[7] was used to impute genotypes from a custom MVP imputation panel including 1000 Genomes phase 3 and enriched for additional African-ancestry genotypes from the African Genome Resource (Sanger Institute) into MVP participants.[8] HARE[3] was used to stratify individuals using a combination of genetically-informed ancestry and self-identified race/ethnicity – we considered individuals of European, African, and Hispanic ancestry. We considered DNA sequence variants with ancestry-specific imputation INFO > 0.3 and minor allele count > 20. Ancestry-specific principal component analysis was performed using EIGENSOFT.[9]

UK Biobank: Details of Pan-UK Biobank genotyping and quality control can be found at https://pan.ukbb.broadinstitute.org/docs/qc and in Bycroft et al.[10] Briefly, 488,377 individuals underwent genotyping from stored blood samples on a custom biobank array (UK BiLEVE or UK biobank Axiom array). Individuals with sex aneuploidy were excluded. Individuals were assigned to continental ancestries based on principal component analysis, projecting UK Biobank individuals into a reference sample of sequence data from 1000 Genomes phase 3 and the Human Genome Diversity Panel. In UK Biobank, we considered individuals of European or African ancestry with INFO scores > 0.8 and minor allele counts > 20.

eMERGE: Genotyping and quality control in eMERGE has been previously reported.[11] Genotyping was performed separately across contributing center-sites using Ilumina and Affymetrix genotype arrays. Imputation was performed using minimac3[7] on the Michigan Imputation Server (https://imputationserver.sph.umich.edu/). Samples with low call-rate (>2% missingness) or duplicated samples were excluded. Continental ancestry was assigned using principal component analysis. We considered individuals of African or European ancestry.

BioBank Japan: Genotyping and quality control in BioBank Japan has been previously reported.[12] Briefly, individuals were genotyped using the Illumina HumanOmniExpressExome BeadChip or a combination of the Illumina HumanOmniExpress and HumanExome BeadChip. Variants were imputed with a custom imputation panel including samples from 1000 Genomes Project phase 3 and enriched with whole-genome sequencing from 1,037 additional Japanese individuals, using minimac3.[7] Following imputation, variants with $r^2<0.7$ were excluded.

FinnGen: Details of genotyping and quality control are available from https://finngen.gitbook.io/documentation/. Briefly, individuals underwent genotyping using Illumina or Affymetrix chip arrays. Individuals with ambiguous gender, high genotype missingness (>5%), excess heterozygosity (+/- 4 standard deviation), and non-Finnish ancestry were excluded. Variants with high-missingness, low Hardy-Weinberg Equilibrium p-value ) $<1 \times 10^{-6}$), and minor allele count <3 were excluded. Samples were pre-phased using Eagle,[13] and imputed to the SISu v3 imputation reference panel using Beagle 4.1.[14] LiftOver was used to map genome positions from hg38 to hg19.[15]

### SUPPLEMENTAL FIGURES

**SUPPLEMENTAL FIGURE 1: Regional Association Plots for Varicose Veins Risk Loci**

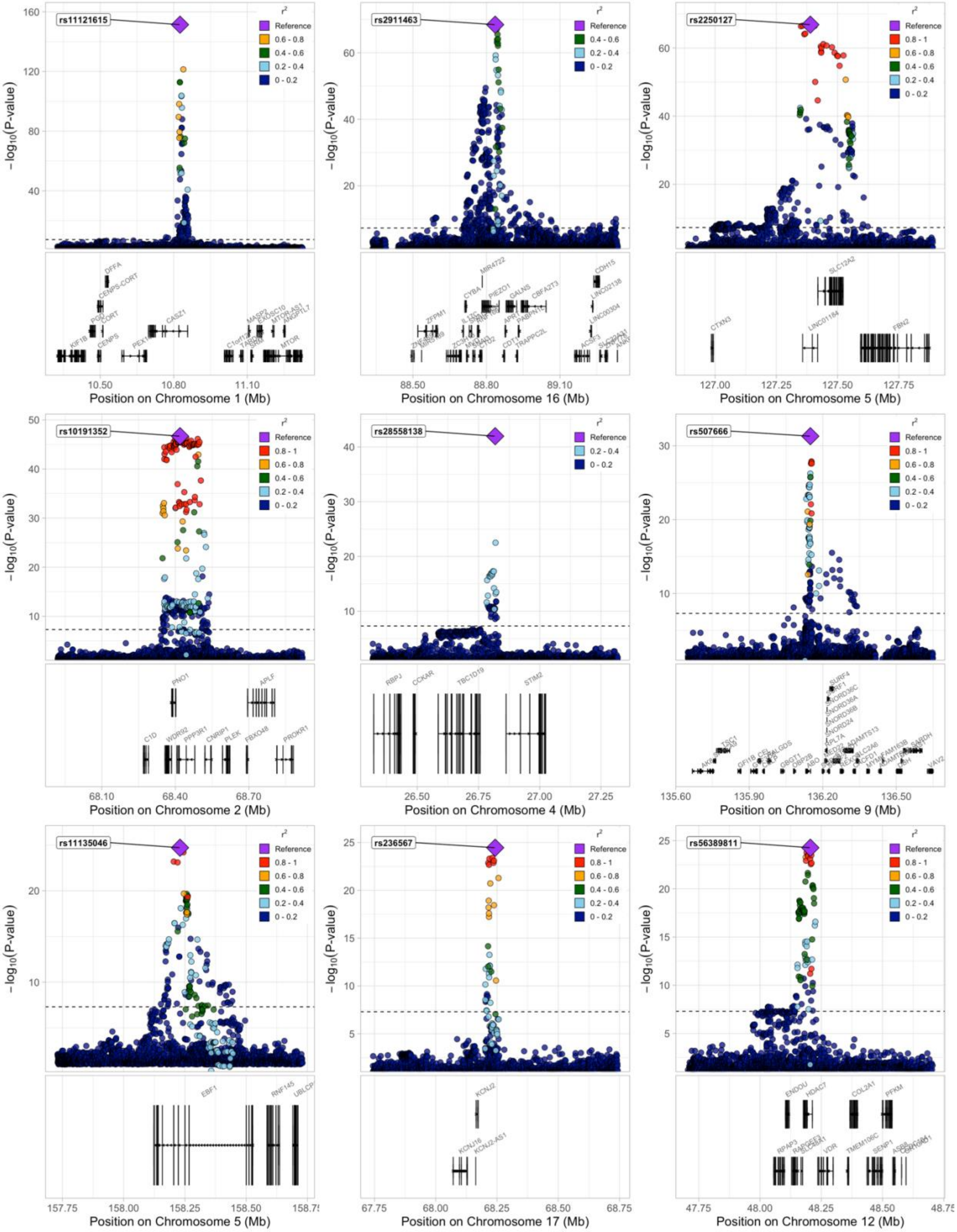

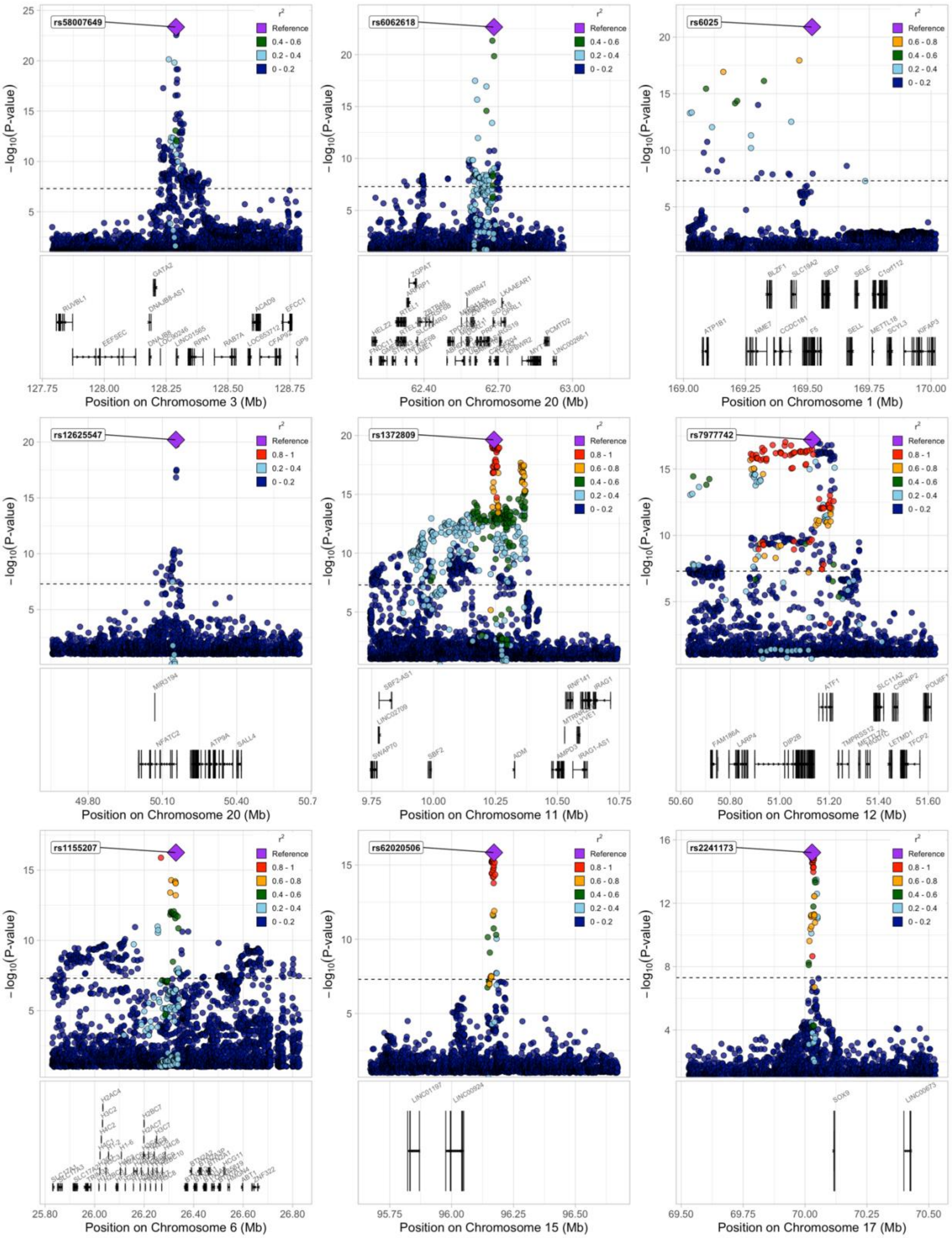
rs58007649
rs6062618
rs6025
rs12625547
rs1372809
rs7977742
rs1155207
rs62020506
rs2241173
$r^2$
Reference
0.8 - 1
0.6 - 0.8
0.4 - 0.6
0.2 - 0.4
0 - 0.2
$-\log_{10}$(P-value)
Position on Chromosome 3 (Mb)
Position on Chromosome 20 (Mb)
Position on Chromosome 1 (Mb)
Position on Chromosome 20 (Mb)
Position on Chromosome 11 (Mb)
Position on Chromosome 12 (Mb)
Position on Chromosome 6 (Mb)
Position on Chromosome 15 (Mb)
Position on Chromosome 17 (Mb)

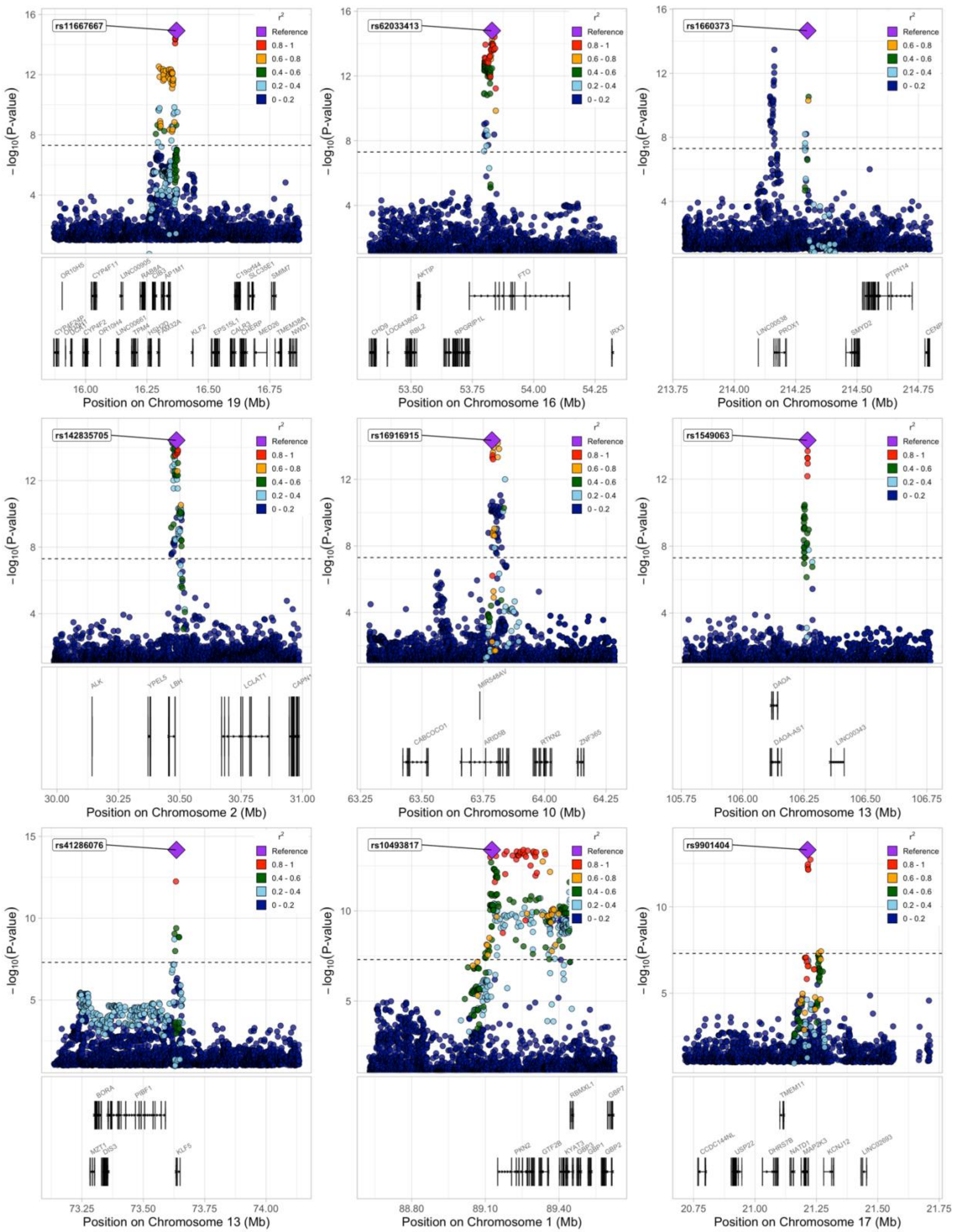
rs11667667
rs62033413
rs1660373
rs142835705
rs16916915
rs1549063
rs41286076
rs10493817
rs9901404
r²
Reference
0.8 - 1
0.6 - 0.8
0.4 - 0.6
0.2 - 0.4
0 - 0.2
−log10(P-value)
Position on Chromosome 19 (Mb)
Position on Chromosome 16 (Mb)
Position on Chromosome 1 (Mb)
Position on Chromosome 2 (Mb)
Position on Chromosome 10 (Mb)
Position on Chromosome 13 (Mb)
Position on Chromosome 13 (Mb)
Position on Chromosome 1 (Mb)
Position on Chromosome 17 (Mb)
AKTIP
FTO
IRX3
PTPN14
PROX1
SMYD2
ALK
YPEL5
LBH
LCLAT1
MIR548AV
ARID5B
RTKN2
ZNF365
DAOA
DAOA-AS1
BORA
PIBF1
DIS3
KLF5
RBMXL1
GBP7
PKN2
GTF2B
GBP1
GBP2
TMEM11
USP22
DHRS7B
NATD1
MAP2K3
KCNJ12

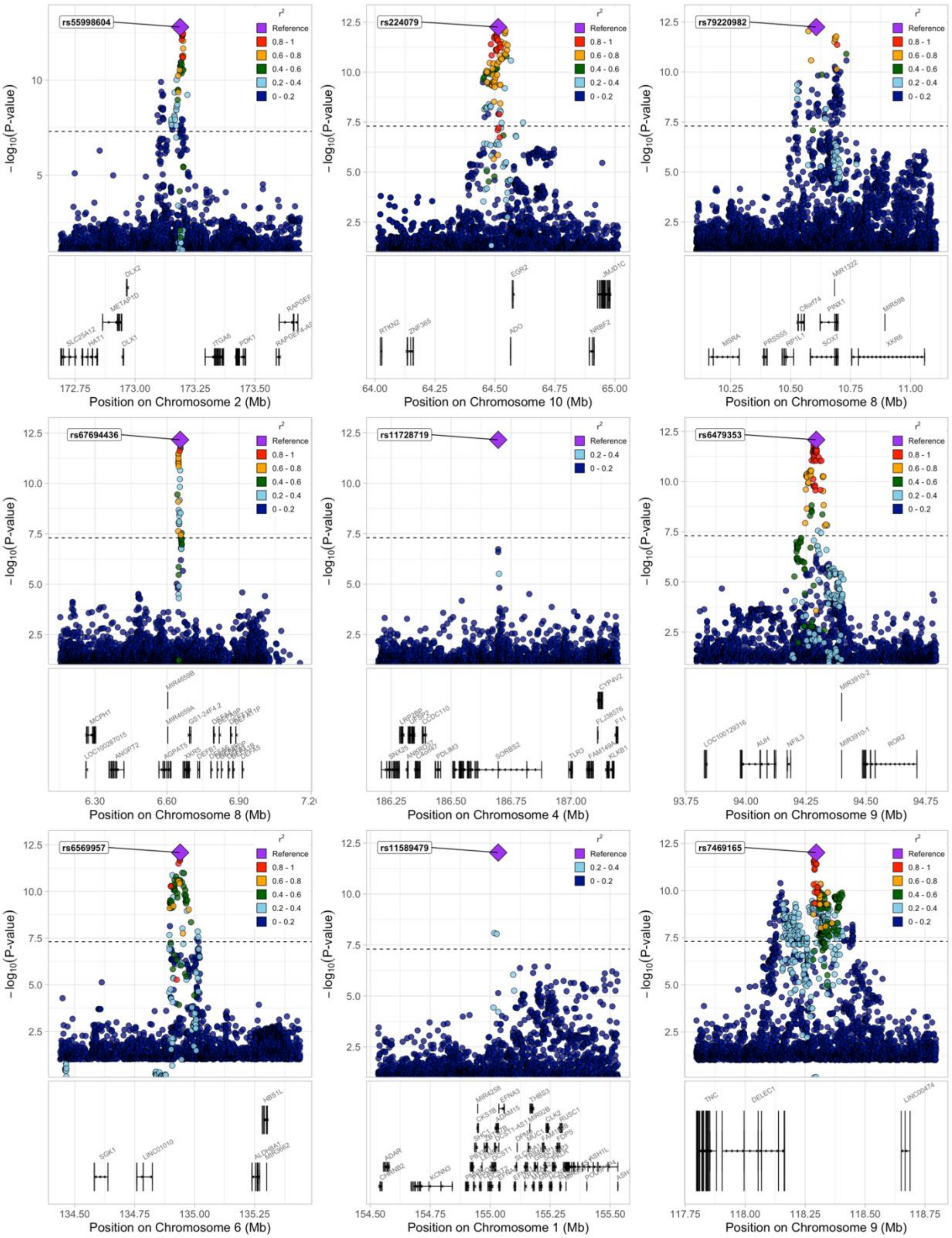
rs55998604
rs224079
rs79220982
rs67694436
rs11728719
rs6479353
rs6569957
rs11589479
rs7469165
r²
Reference
0.8 - 1
0.6 - 0.8
0.4 - 0.6
0.2 - 0.4
0 - 0.2
−log₁₀(P-value)
Position on Chromosome 2 (Mb)
Position on Chromosome 10 (Mb)
Position on Chromosome 8 (Mb)
Position on Chromosome 8 (Mb)
Position on Chromosome 4 (Mb)
Position on Chromosome 9 (Mb)
Position on Chromosome 6 (Mb)
Position on Chromosome 1 (Mb)
Position on Chromosome 9 (Mb)

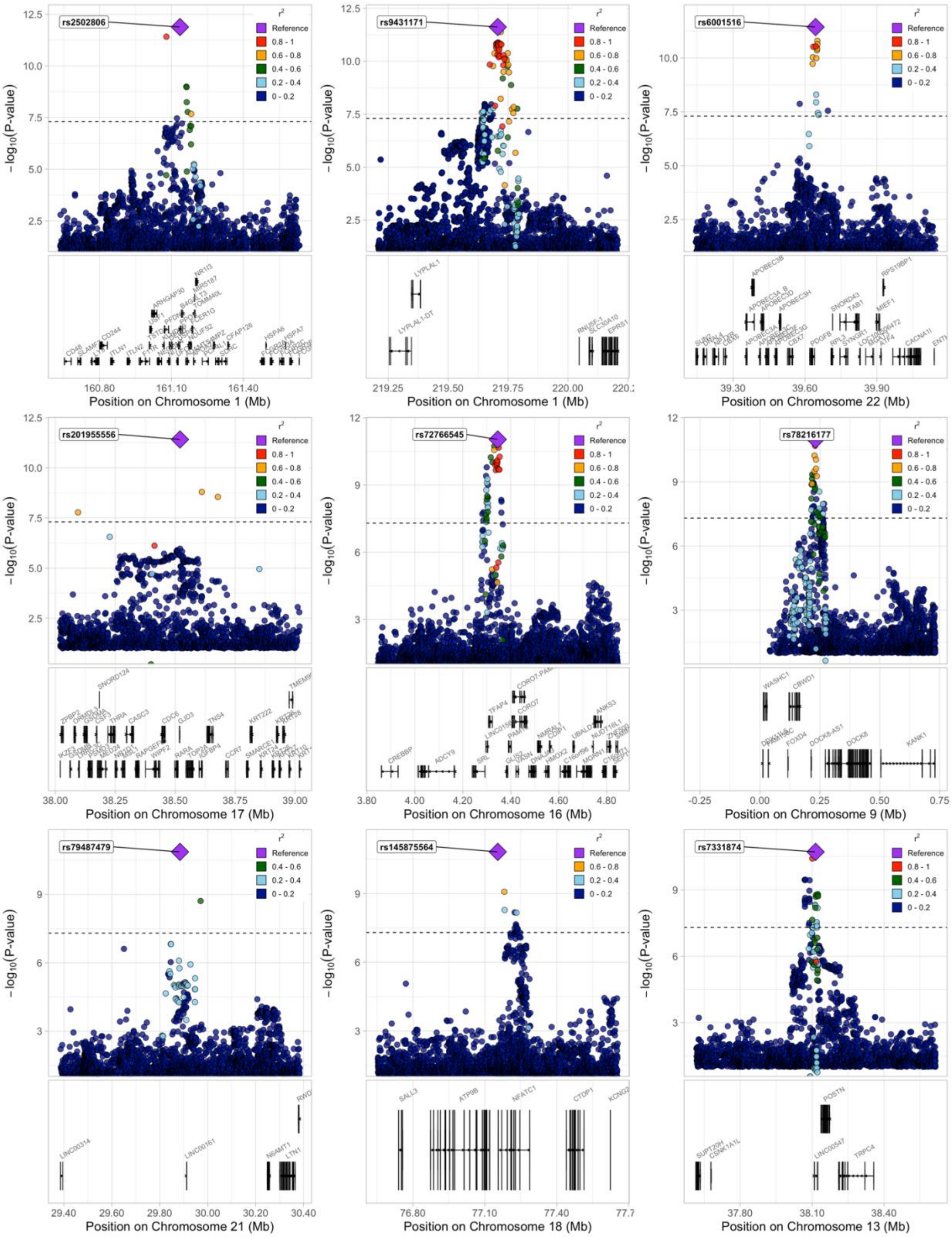
rs2502806
rs9431171
rs6001516
rs201955556
rs72766545
rs78216177
rs79487479
rs145875564
rs7331874
$-\log_{10}$(P-value)
$r^2$
Reference
0.8 - 1
0.6 - 0.8
0.4 - 0.6
0.2 - 0.4
0 - 0.2
Position on Chromosome 1 (Mb)
Position on Chromosome 1 (Mb)
Position on Chromosome 22 (Mb)
Position on Chromosome 17 (Mb)
Position on Chromosome 16 (Mb)
Position on Chromosome 9 (Mb)
Position on Chromosome 21 (Mb)
Position on Chromosome 18 (Mb)
Position on Chromosome 13 (Mb)
LYPLAL1
LYPLAL1-DT
APOBEC3B
SNORD124
WASHC1
CBWD1
DOCK8
KANK1
LINC00314
LINC00161
SALL3
ATP9B
NFATC1
CTDP1
POSTN
TRPC4

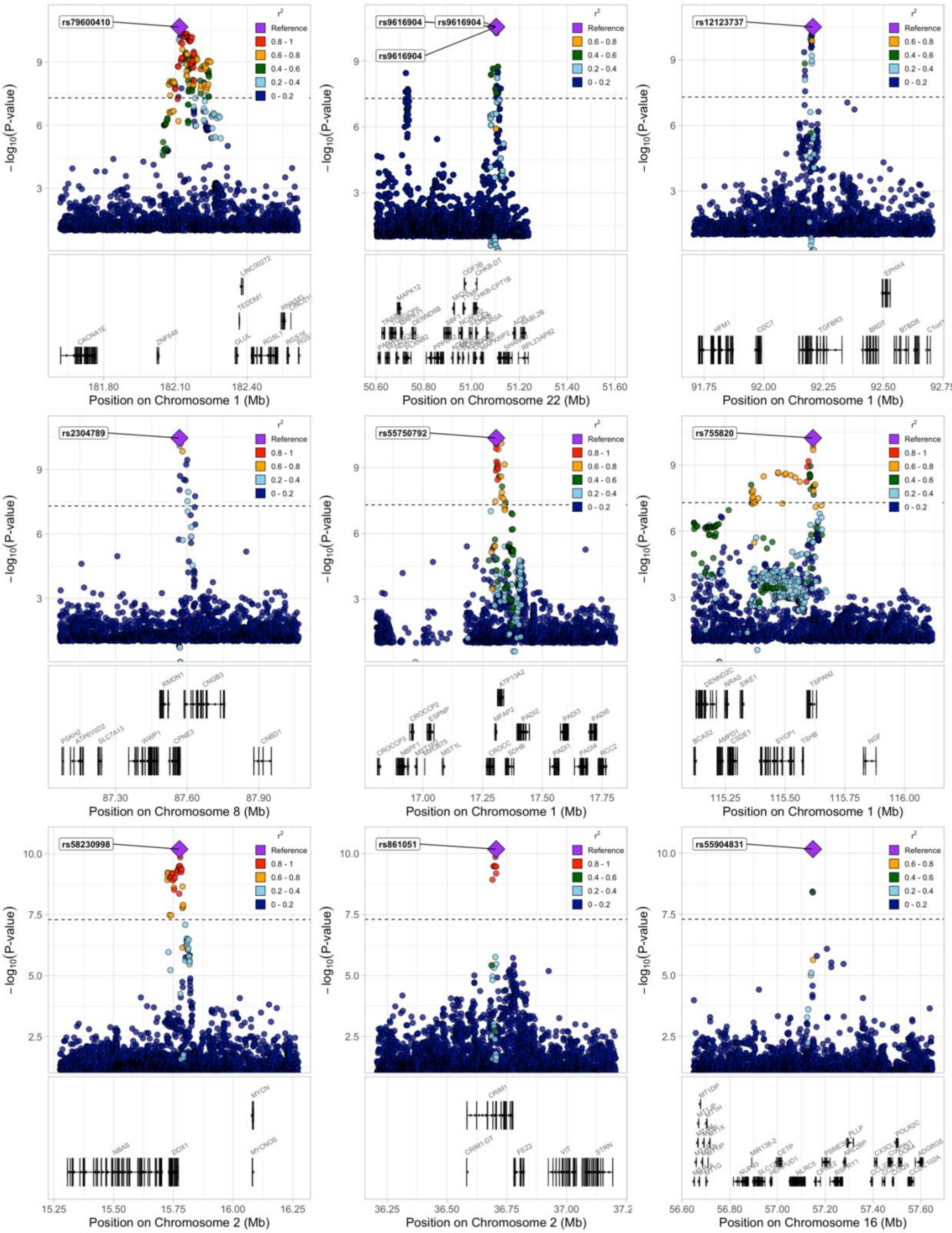
rs79600410
rs9616904
rs12123737
rs2304789
rs55750792
rs755820
rs58230998
rs861051
rs55904831
r²
Reference
0.8 - 1
0.6 - 0.8
0.4 - 0.6
0.2 - 0.4
0 - 0.2
−log10(P-value)
Position on Chromosome 1 (Mb)
Position on Chromosome 22 (Mb)
Position on Chromosome 8 (Mb)
Position on Chromosome 2 (Mb)
Position on Chromosome 16 (Mb)

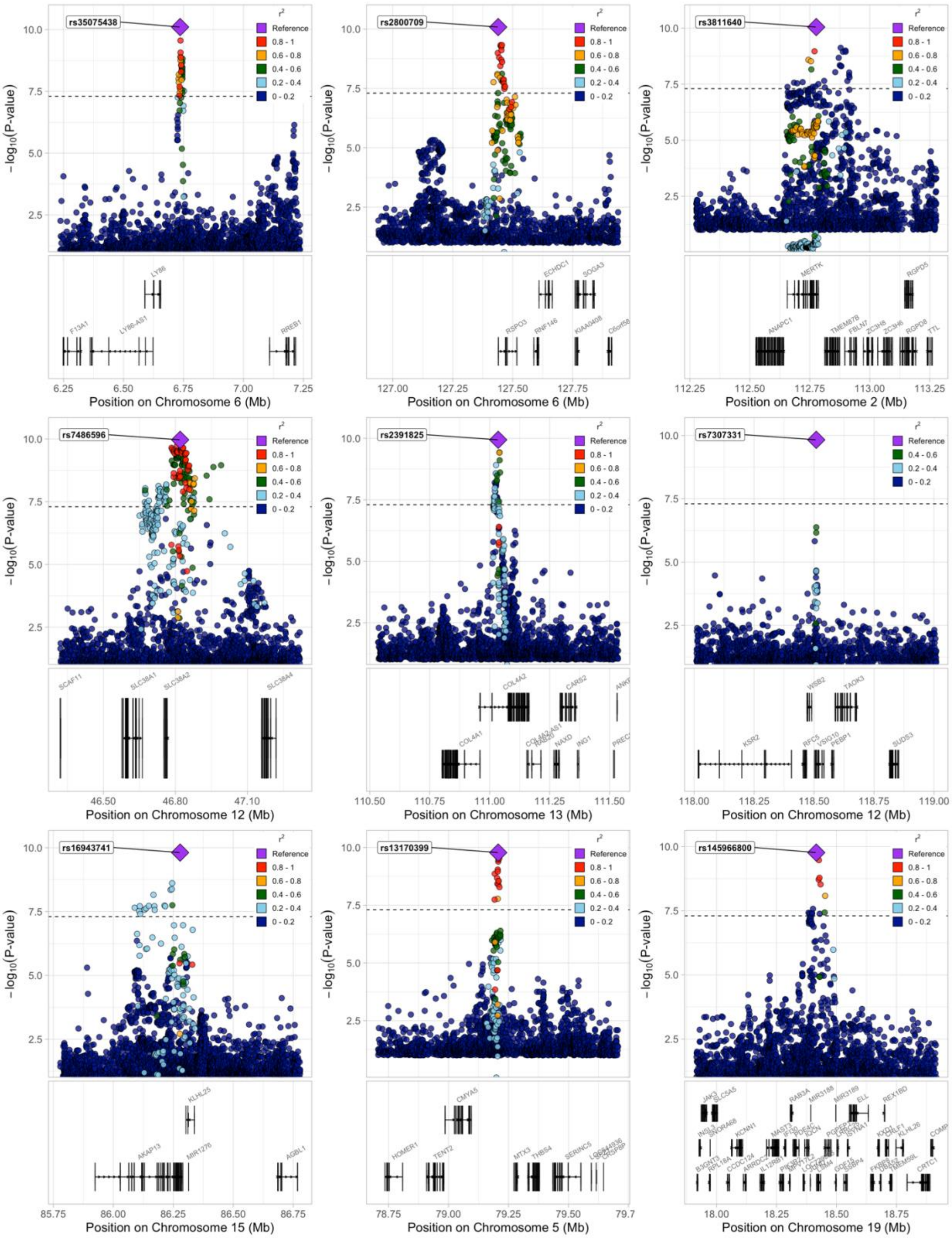
rs35075438
rs2800709
rs3811640
rs7486596
rs2391825
rs7307331
rs16943741
rs13170399
rs145966800
$r^2$
Reference
0.8 - 1
0.6 - 0.8
0.4 - 0.6
0.2 - 0.4
0 - 0.2
$-\log_{10}$(P-value)
Position on Chromosome 6 (Mb)
Position on Chromosome 6 (Mb)
Position on Chromosome 2 (Mb)
Position on Chromosome 12 (Mb)
Position on Chromosome 13 (Mb)
Position on Chromosome 12 (Mb)
Position on Chromosome 15 (Mb)
Position on Chromosome 5 (Mb)
Position on Chromosome 19 (Mb)

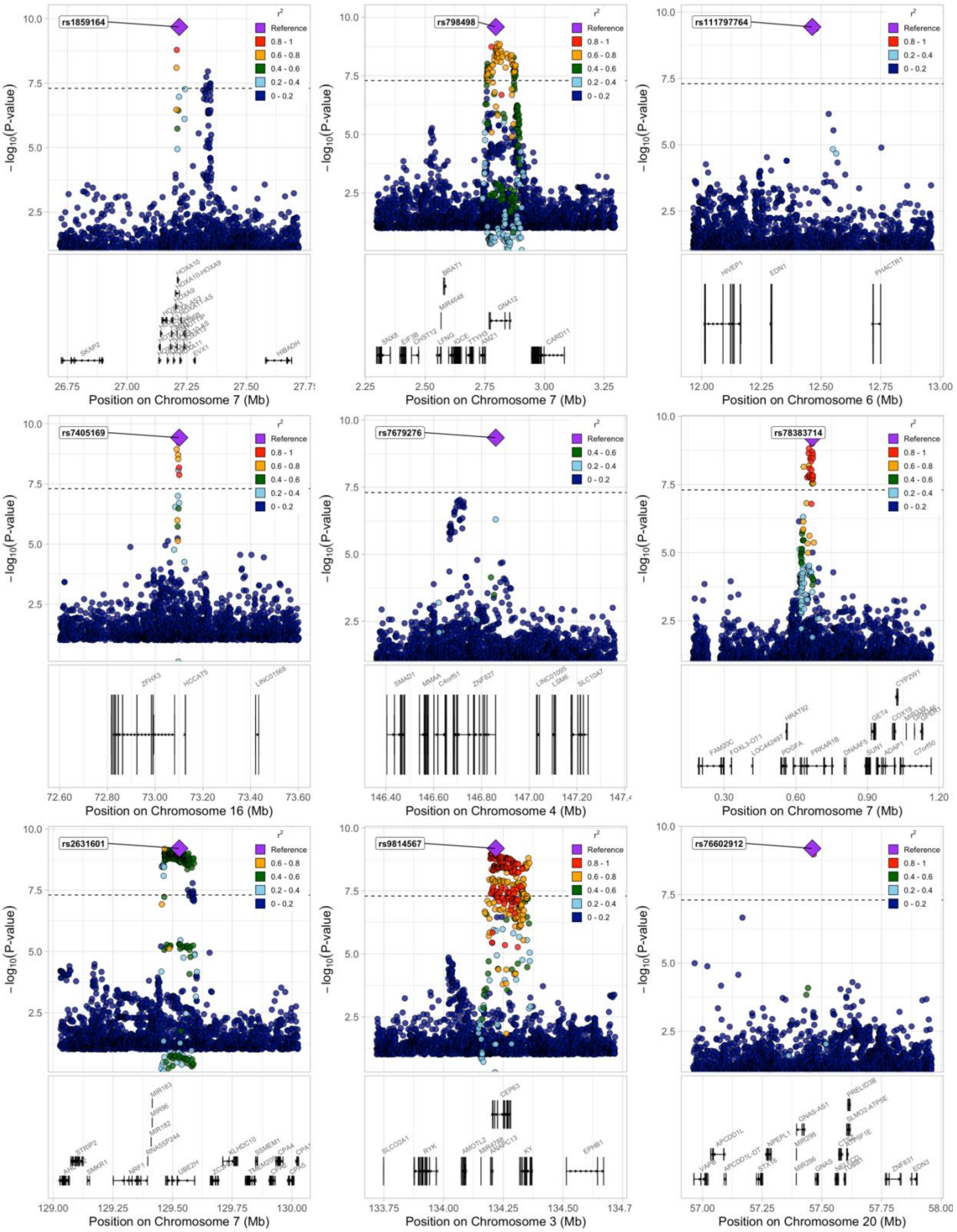
rs1859164
rs798498
rs111797764
rs7405169
rs7679276
rs78383714
rs2631601
rs9814567
rs76602912
$r^2$
Reference
0.8 - 1
0.6 - 0.8
0.4 - 0.6
0.2 - 0.4
0 - 0.2
$-\log_{10}$(P-value)
Position on Chromosome 7 (Mb)
Position on Chromosome 6 (Mb)
Position on Chromosome 16 (Mb)
Position on Chromosome 4 (Mb)
Position on Chromosome 3 (Mb)
Position on Chromosome 20 (Mb)

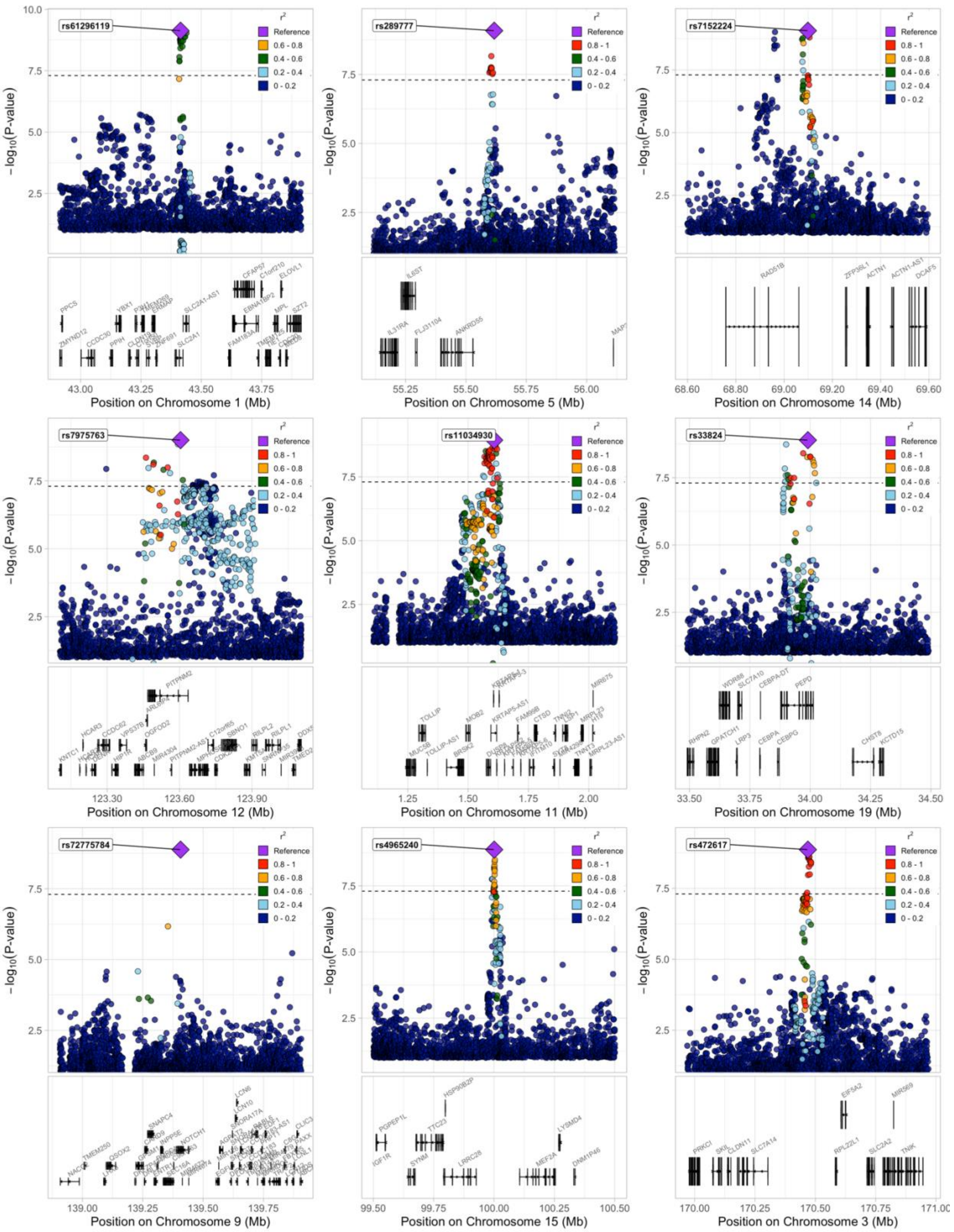

rs61296119
rs289777
rs7152224
rs7975763
rs11034930
rs33824
rs72775784
rs4965240
rs472617
$-\log_{10}$(P-value)
$r^2$
Reference
0.8 - 1
0.6 - 0.8
0.4 - 0.6
0.2 - 0.4
0 - 0.2
Position on Chromosome 1 (Mb)
Position on Chromosome 5 (Mb)
Position on Chromosome 14 (Mb)
Position on Chromosome 12 (Mb)
Position on Chromosome 11 (Mb)
Position on Chromosome 19 (Mb)
Position on Chromosome 9 (Mb)
Position on Chromosome 15 (Mb)
Position on Chromosome 3 (Mb)

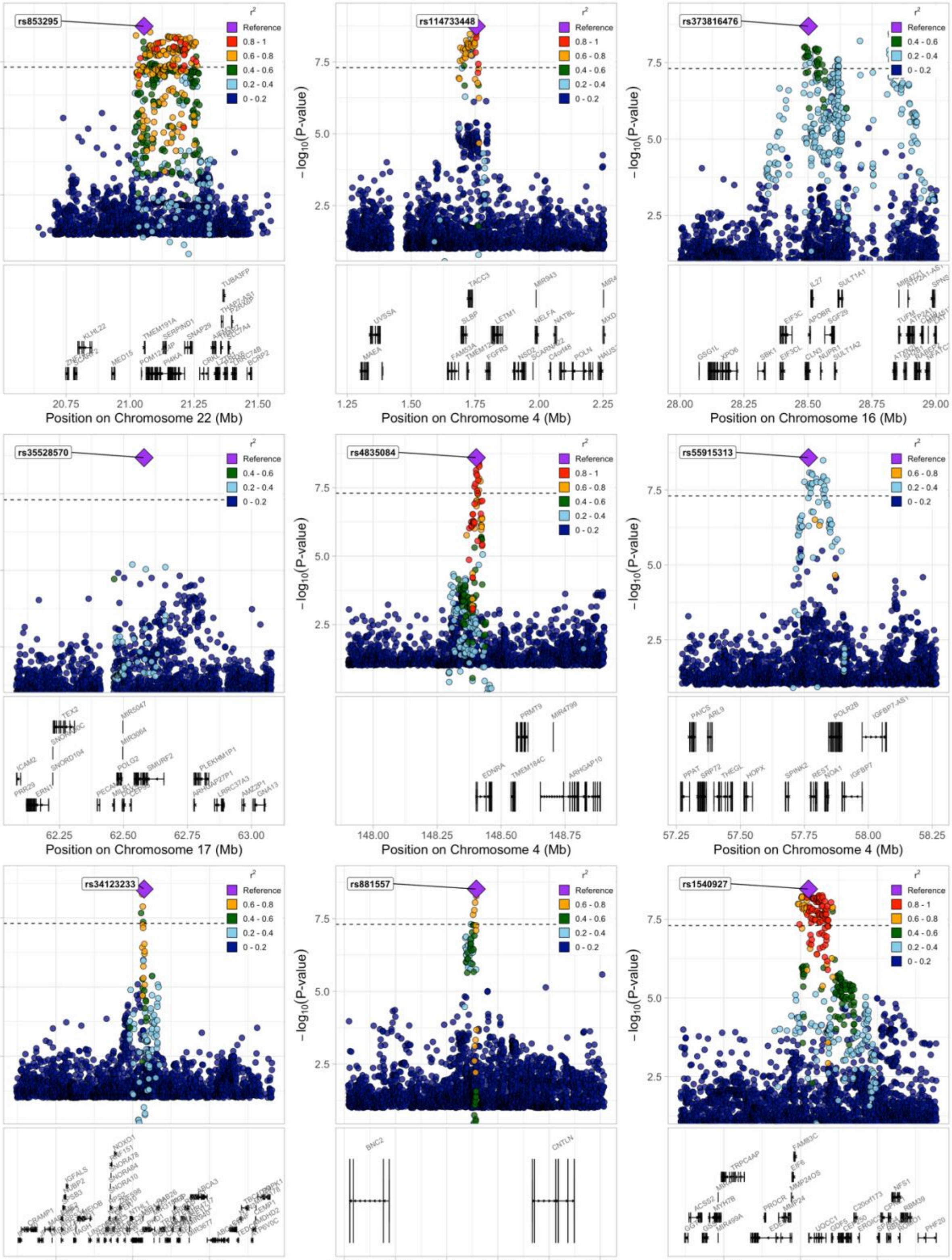
rs853295
rs114733448
rs373816476
rs35528570
rs4835084
rs55915313
rs34123233
rs881557
rs1540927
−log10(P-value)
r²
Reference
0.8 - 1
0.6 - 0.8
0.4 - 0.6
0.2 - 0.4
0 - 0.2
Position on Chromosome 22 (Mb)
Position on Chromosome 4 (Mb)
Position on Chromosome 16 (Mb)
Position on Chromosome 17 (Mb)
Position on Chromosome 4 (Mb)
Position on Chromosome 4 (Mb)
Position on Chromosome 16 (Mb)
Position on Chromosome 9 (Mb)
Position on Chromosome 20 (Mb)

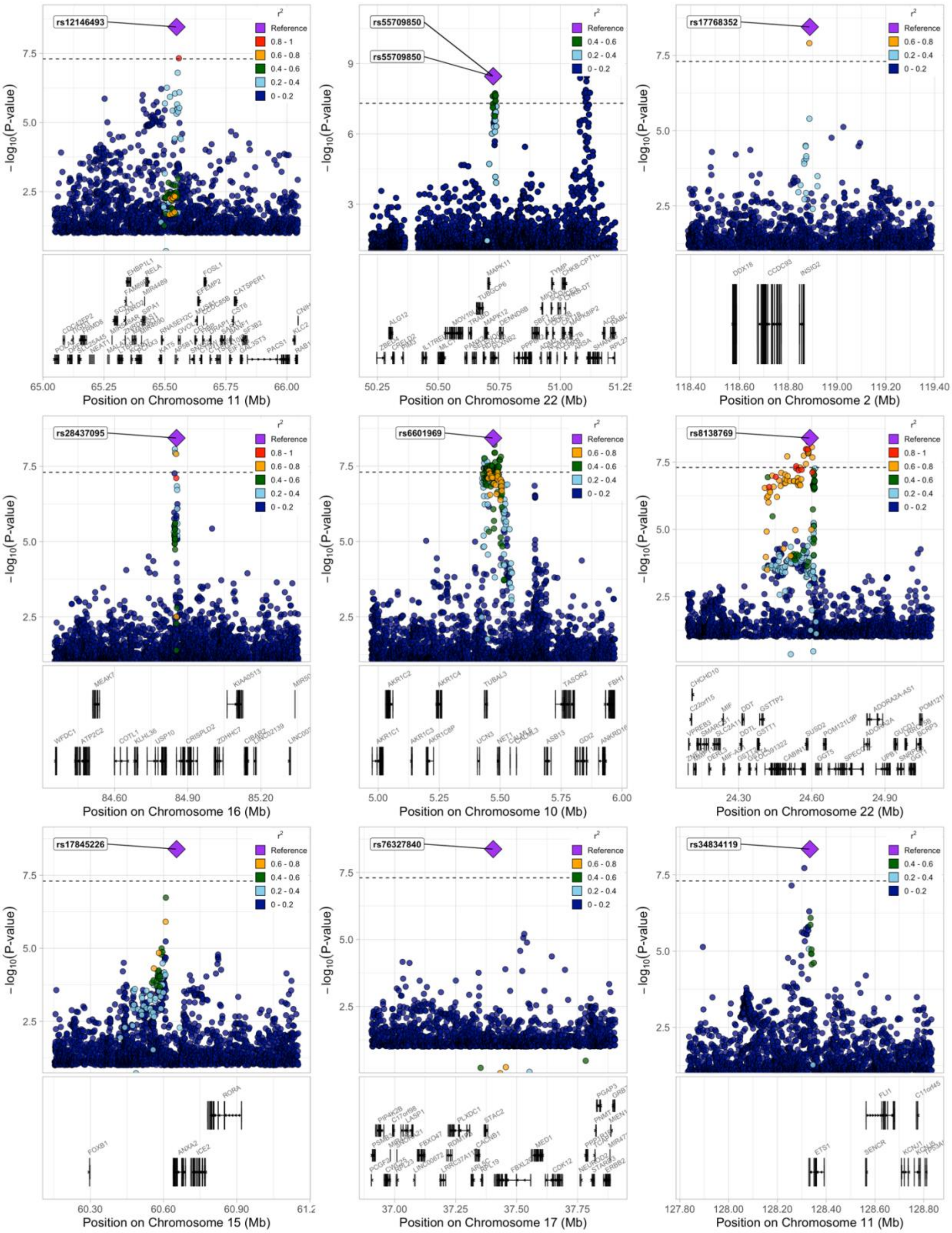
rs12146493
rs55709850
rs55709850
rs17768352
rs28437095
rs6601969
rs8138769
rs17845226
rs76327840
rs34834119
r²
Reference
0.8 - 1
0.6 - 0.8
0.4 - 0.6
0.2 - 0.4
0 - 0.2
-log10(P-value)
Position on Chromosome 11 (Mb)
Position on Chromosome 22 (Mb)
Position on Chromosome 2 (Mb)
Position on Chromosome 16 (Mb)
Position on Chromosome 10 (Mb)
Position on Chromosome 22 (Mb)
Position on Chromosome 15 (Mb)
Position on Chromosome 17 (Mb)
Position on Chromosome 11 (Mb)

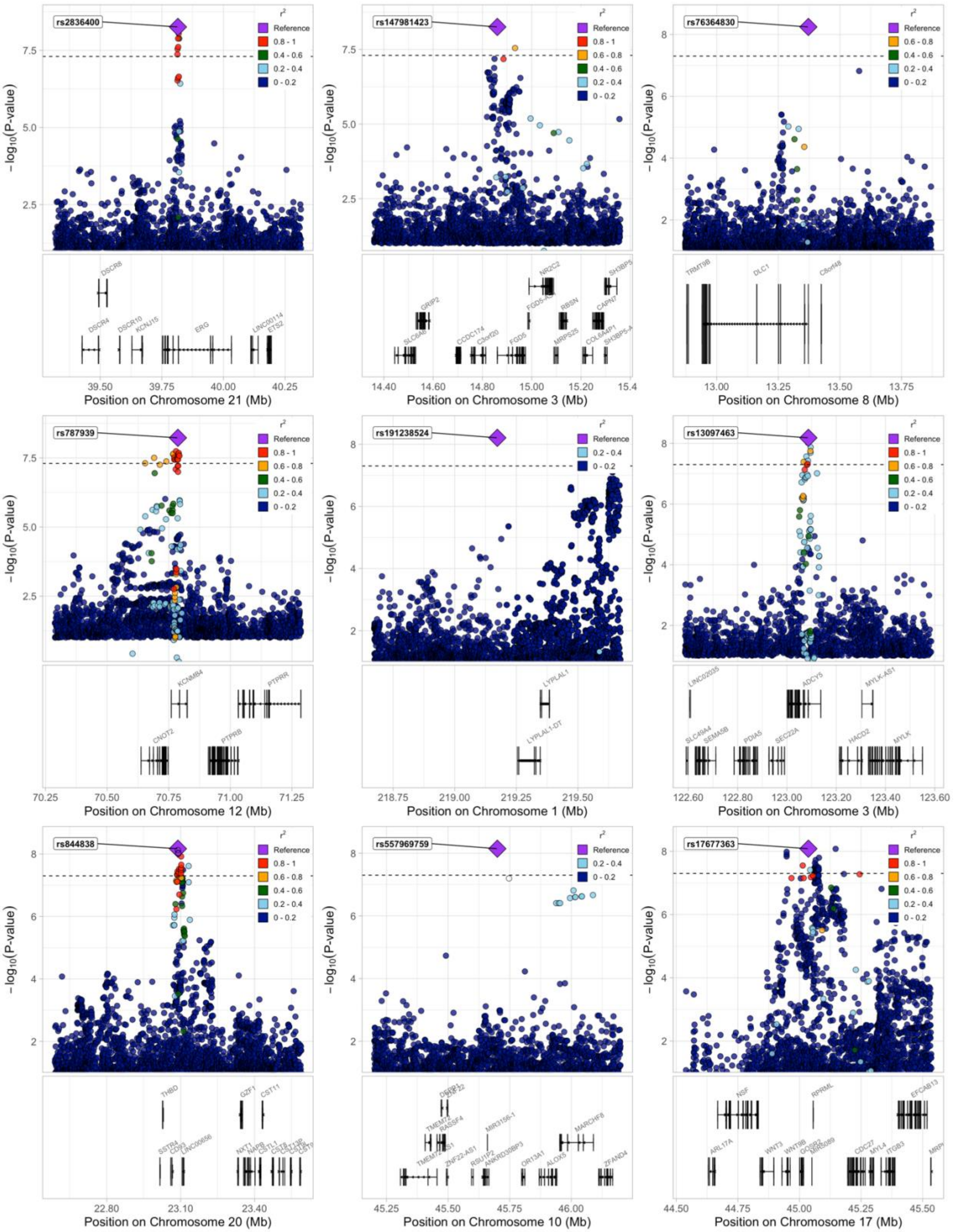
rs2836400
rs147981423
rs76364830
rs787939
rs191238524
rs13097463
rs844838
rs557969759
rs17677363
r²
Reference
0.8 - 1
0.6 - 0.8
0.4 - 0.6
0.2 - 0.4
0 - 0.2
−log10(P-value)
Position on Chromosome 21 (Mb)
Position on Chromosome 3 (Mb)
Position on Chromosome 8 (Mb)
Position on Chromosome 12 (Mb)
Position on Chromosome 1 (Mb)
Position on Chromosome 3 (Mb)
Position on Chromosome 20 (Mb)
Position on Chromosome 10 (Mb)
Position on Chromosome 17 (Mb)

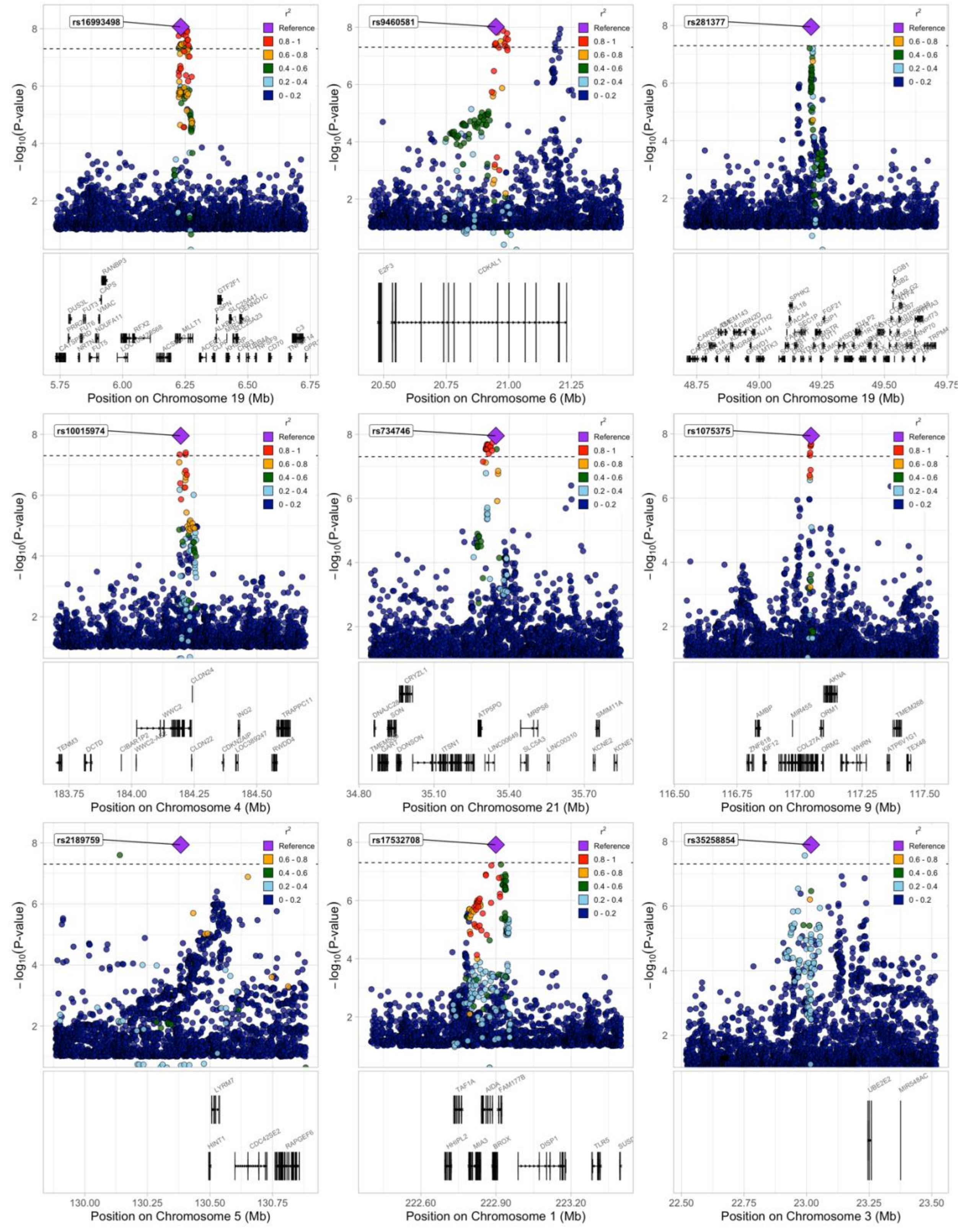

rs16993498
rs9460581
rs281377
rs10015974
rs734746
rs1075375
rs2189759
rs17532708
rs35258854
r²
Reference
0.8 - 1
0.6 - 0.8
0.4 - 0.6
0.2 - 0.4
0 - 0.2
−log₁₀(P-value)
Position on Chromosome 19 (Mb)
Position on Chromosome 6 (Mb)
Position on Chromosome 19 (Mb)
Position on Chromosome 4 (Mb)
Position on Chromosome 21 (Mb)
Position on Chromosome 9 (Mb)
Position on Chromosome 5 (Mb)
Position on Chromosome 1 (Mb)
Position on Chromosome 3 (Mb)

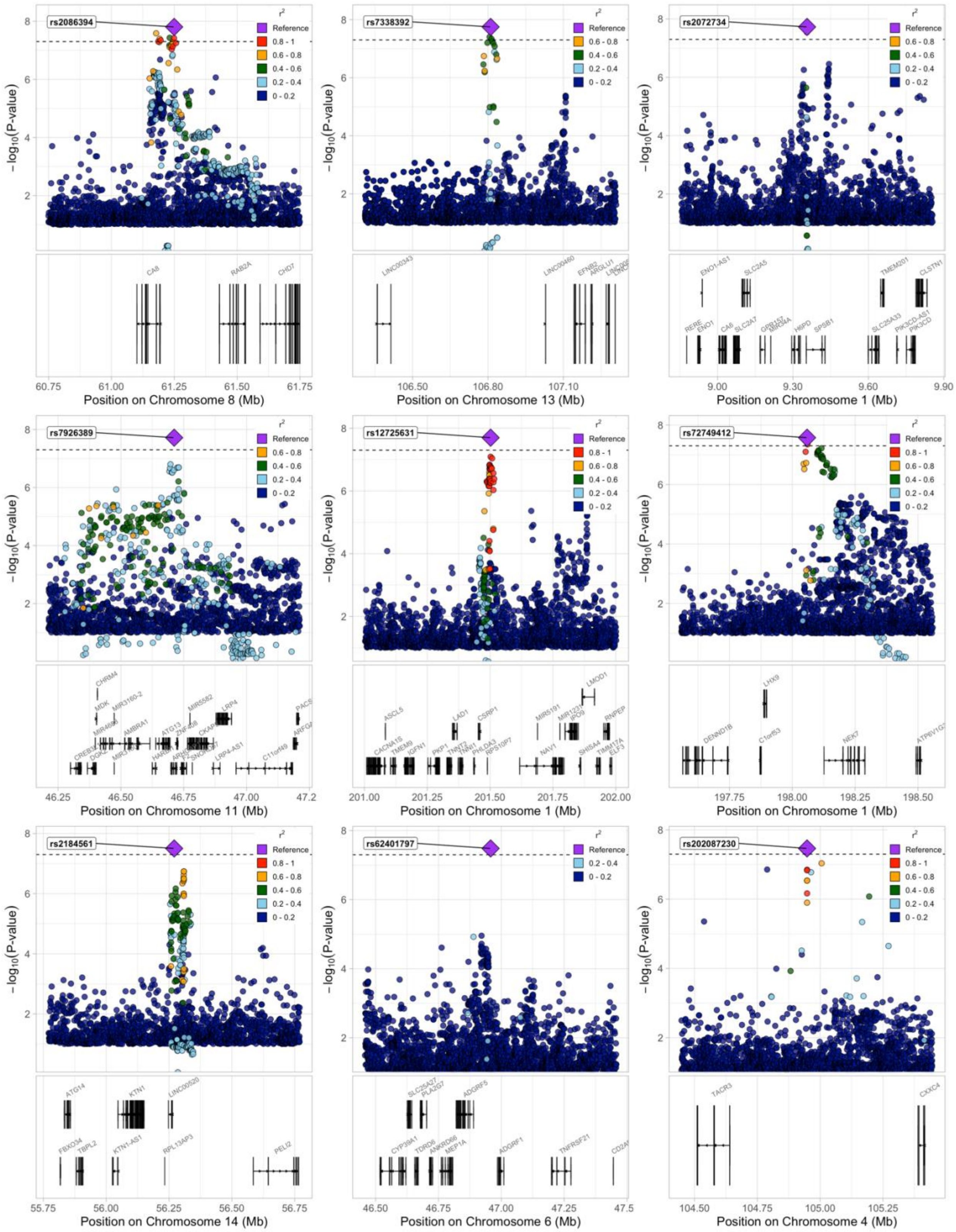
rs2086394
rs7338392
rs2072734
rs7926389
rs12725631
rs72749412
rs2184561
rs62401797
rs202087230
r2
Reference
0.8 - 1
0.6 - 0.8
0.4 - 0.6
0.2 - 0.4
0 - 0.2
−log10(P-value)
Position on Chromosome 8 (Mb)
Position on Chromosome 13 (Mb)
Position on Chromosome 1 (Mb)
Position on Chromosome 11 (Mb)
Position on Chromosome 1 (Mb)
Position on Chromosome 1 (Mb)
Position on Chromosome 14 (Mb)
Position on Chromosome 6 (Mb)
Position on Chromosome 4 (Mb)

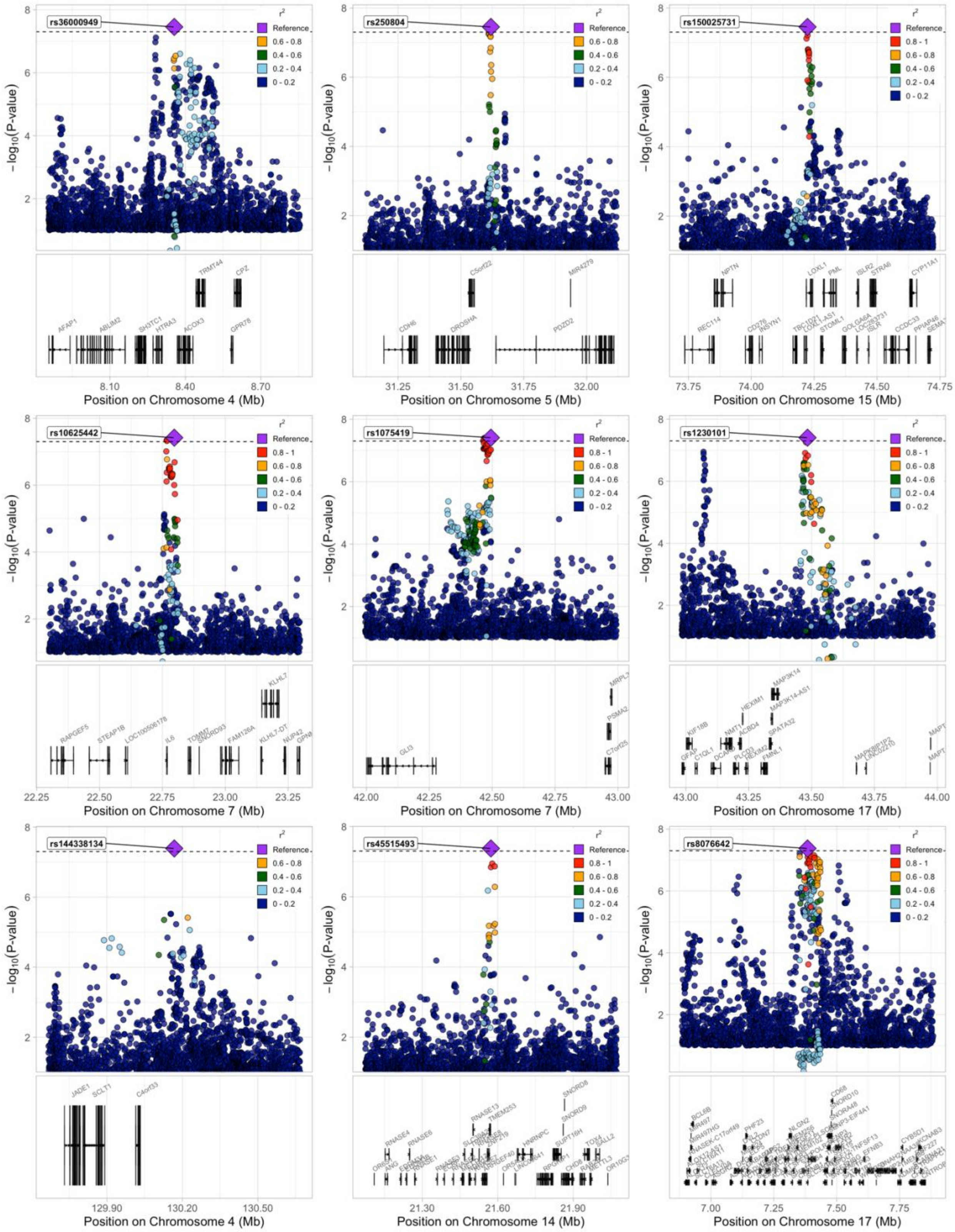
rs36000949
rs250804
rs150025731
rs10625442
rs1075419
rs1230101
rs144338134
rs45515493
rs8076642
$-\log_{10}$(P-value)
$r^2$
Reference
0.8 - 1
0.6 - 0.8
0.4 - 0.6
0.2 - 0.4
0 - 0.2
Position on Chromosome 4 (Mb)
Position on Chromosome 5 (Mb)
Position on Chromosome 15 (Mb)
Position on Chromosome 7 (Mb)
Position on Chromosome 7 (Mb)
Position on Chromosome 17 (Mb)
Position on Chromosome 4 (Mb)
Position on Chromosome 14 (Mb)
Position on Chromosome 17 (Mb)

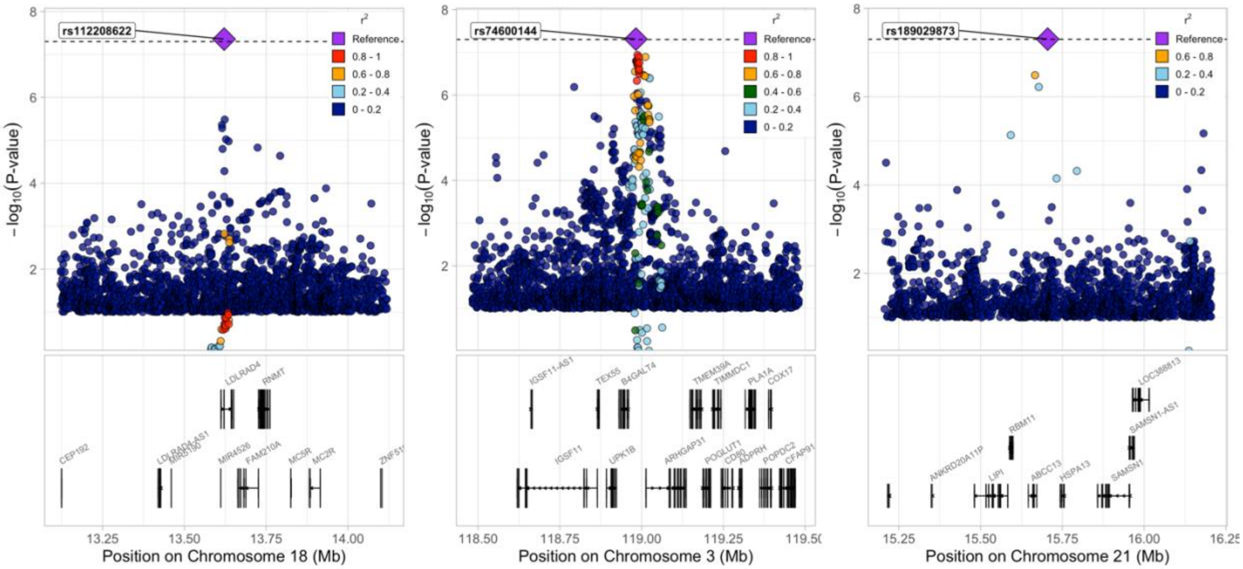
rs112208622
rs74600144
rs189029873
r²
Reference
0.8 - 1
0.6 - 0.8
0.4 - 0.6
0.2 - 0.4
0 - 0.2
−log10(P-value)
LDLRAD4
RNMT
CEP192
MIR4526
FAM210A
MC5R
MC2R
IGSF11-AS1
TEX55
B4GALT4
TMEM39A
TIMMDC1
PLA1A
COX17
IGSF11
UPK1B
ARHGAP31
POGLUT1
CD80
ADPRH
POPDC2
LOC388813
RBM11
SAMSN1-AS1
ANKRD20A11P
LIPI
ABCC13
HSPA13
SAMSN1
Position on Chromosome 18 (Mb)
Position on Chromosome 3 (Mb)
Position on Chromosome 21 (Mb)

**SUPPLEMENTAL FIGURE 2: Effect Size and Minor Allele Frequency for Novel and Previously Reported Risk Loci**

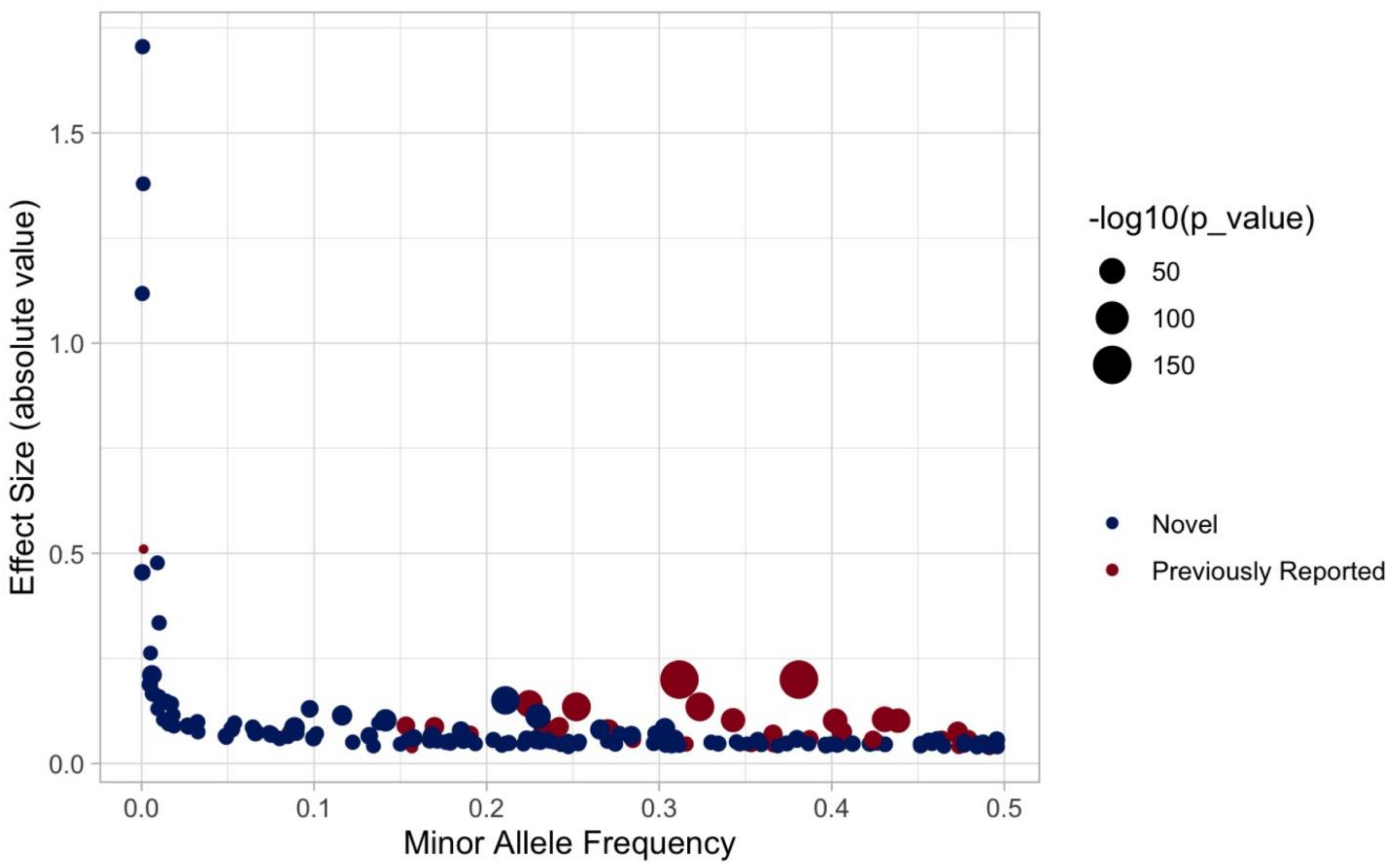

Absolute effect size plotted as a function of minor allele frequency for all novel genome-wide significant loci, and the 30/36 previously reported loci with $p < 0.05$ in the current meta-analysis.

**SUPPLEMENTAL FIGURE 3: MAGMA Tissue and Gene Set Enrichment**

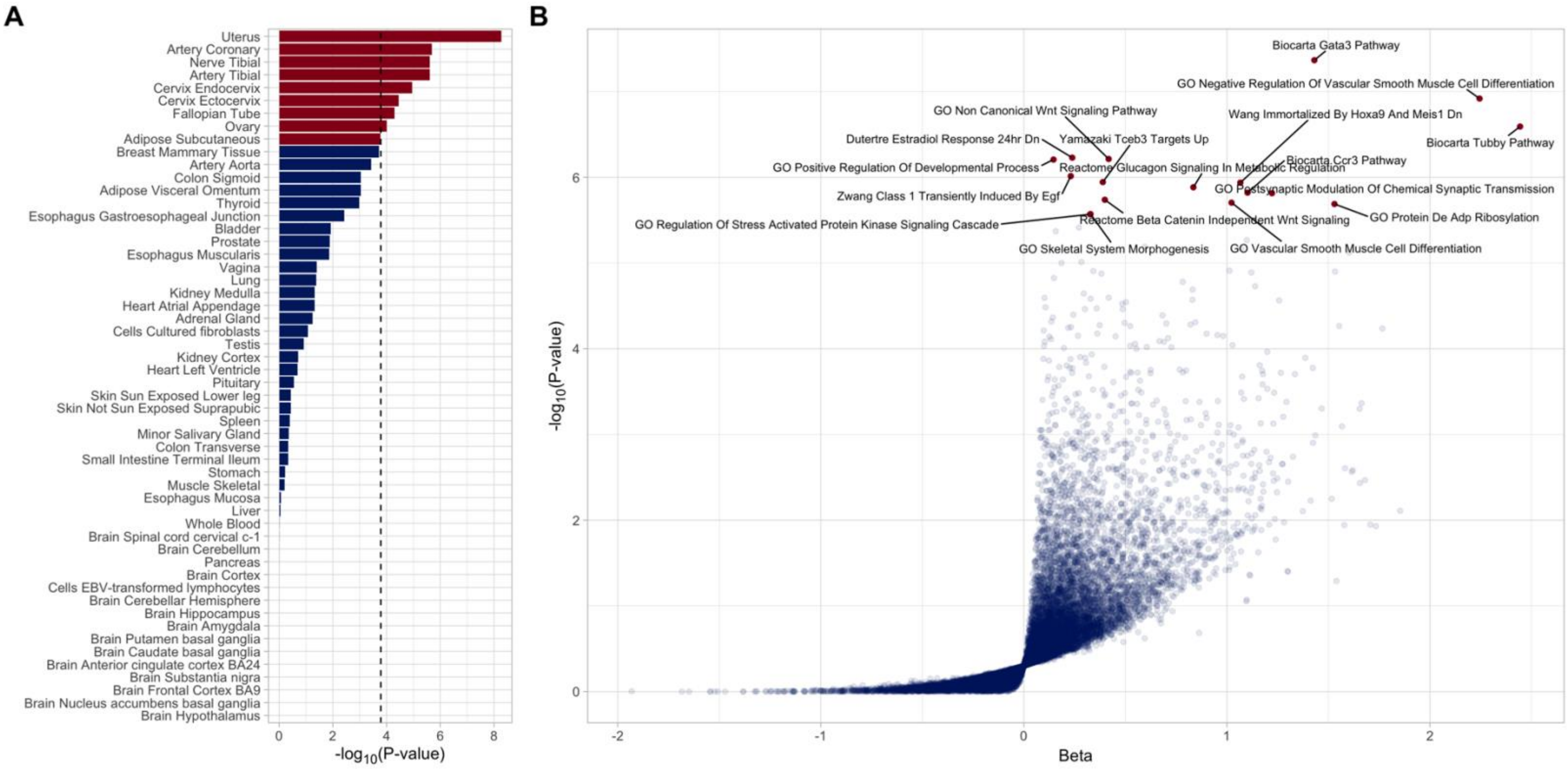

A) Tissue enrichment was assessed using MAGMA via the online FUMA platform, integrating GWAS summary statistics for varicose veins with gene expression (RNAseq) data from GTEx V8. B) Gene set enrichment results from MAGMA.

**SUPPLEMENTAL FIGURE 4: Genetic Correlation Between Varicose Veins and Vascular Traits**

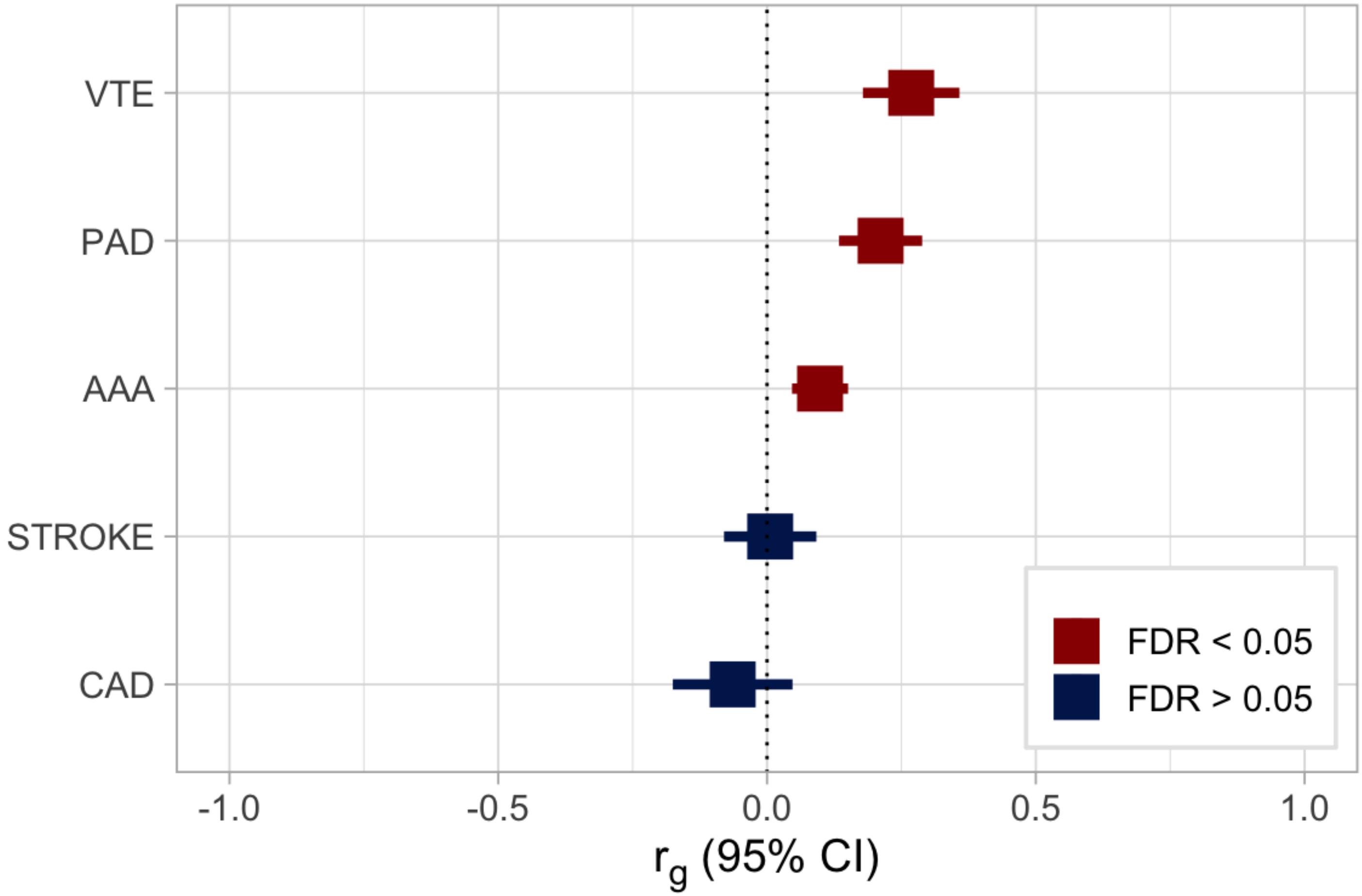

Genetic correlations between varicose veins and other vascular traits were estimated using cross-trait LDSC. Varicose veins were significantly correlated with VTE (venous thromboembolism), PAD (peripheral artery disease), and AAA (abdominal aortic aneurysm), but not STROKE (any ischemic stroke) or CAD (coronary artery disease).

**SUPPLEMENTAL FIGURE 5: Multi-Trait Colocalization**

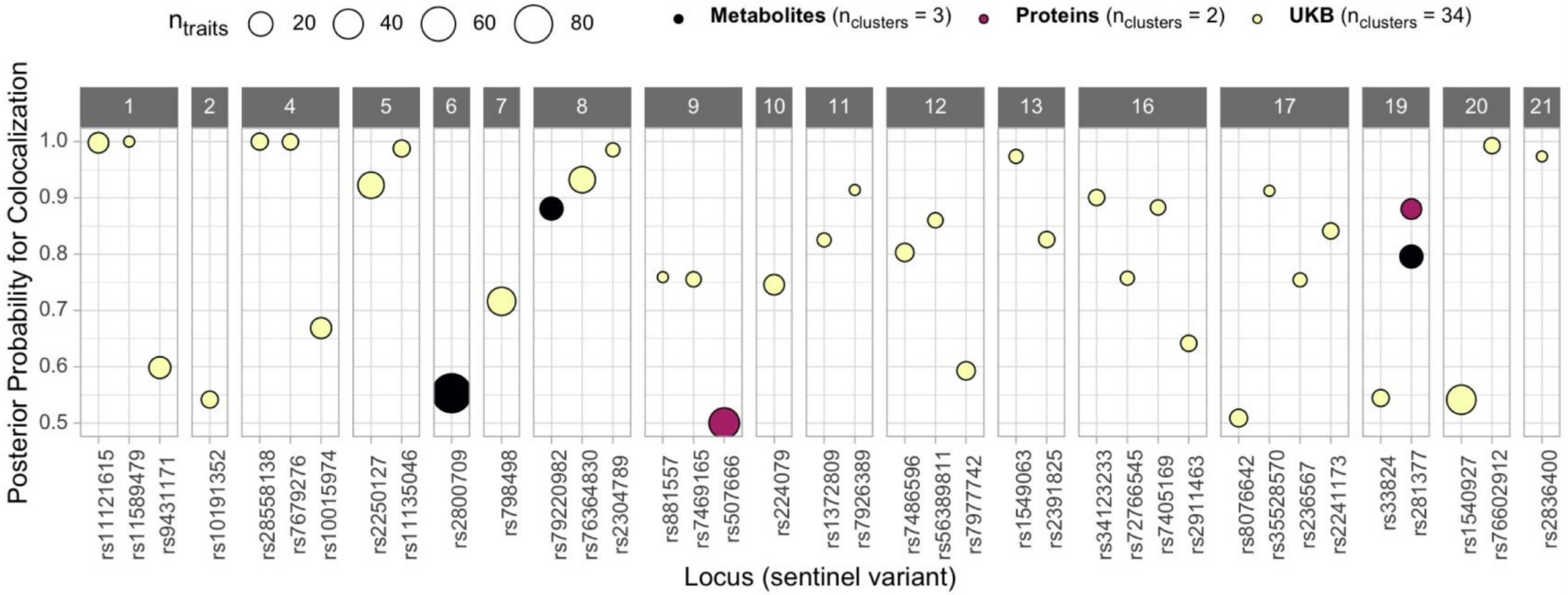

Multi-trait colocalization was performed across all metabolites, proteins, or UK Biobank traits with shared associations with varicose veins at a given locus. Loci are labeled by chromosome, and each locus is denoted by the sentinel variant from the varicose veins GWAS meta-analysis. The size of points corresponds to the number of colocalizing traits.

**SUPPLEMENTAL FIGURE 6: Random Walk with Restart-based Topological Gene Enrichment of Genome-wide Significant Varicose Veins Variants**

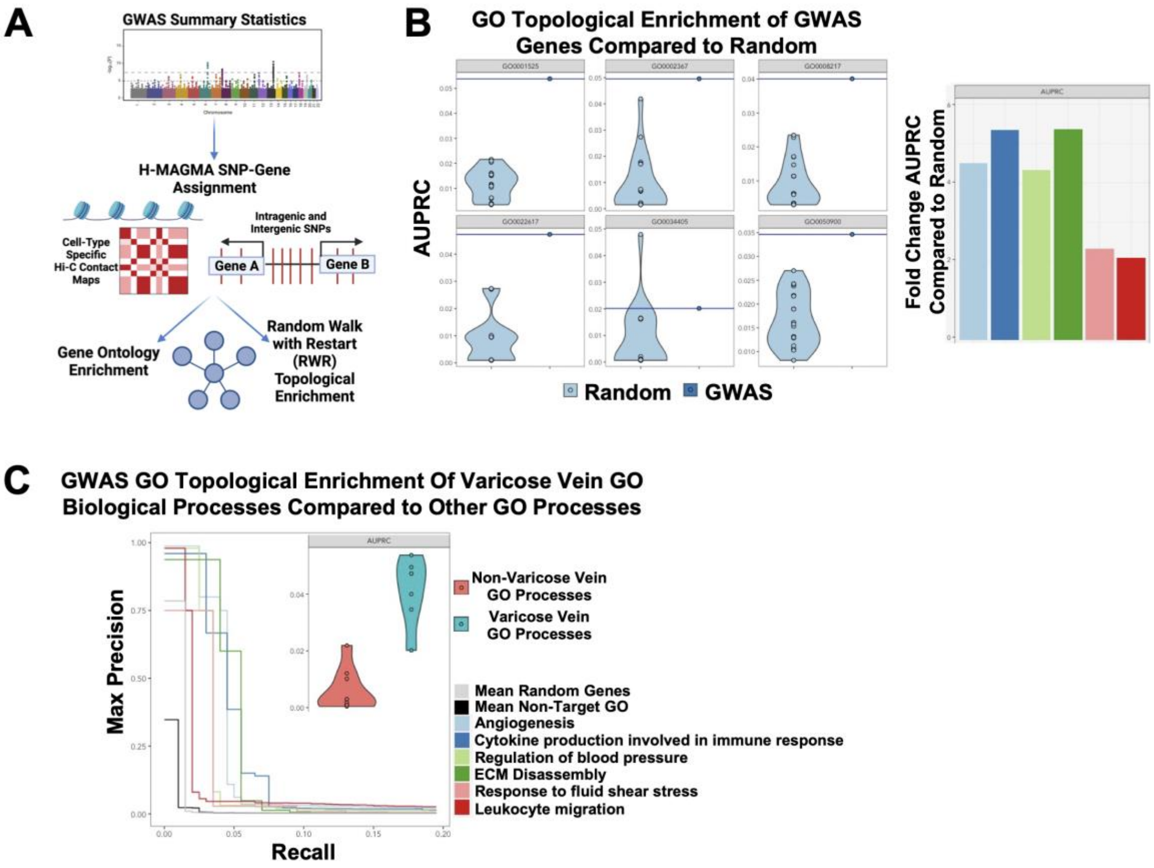

**A)** Workflow of cell-type specific SNP-to-gene assignment and gene set enrichment analysis. **B)** Left: Varicose veins GWAS genes are topologically enriched for GO terms related to known varicose veins pathophysiological processes compared to random gene sets of equivalent size based on random walk with restart (RWR) ranking of genes in GO term. Right: Fold change of AUPRC values for varicose vein GO processes compared to values of random genes. **C)** GWAS genes are more topologically enriched in varicose vein GO processes compared to random genes (gray) and non-varicose vein-related GO processes (black) by interpolated precision values. Inset: Violin plot of AUPRC values for each gene set related to non-target (red) and target (blue) GO biological processes. AUPRC: area under precision-recall curve; ECM: extracellular matrix; Angiogenesis: GO:0001525; Cytokine production involved in immune response: GO:0002367; Regulation of blood pressure: GO:0008217; ECM Disassembly: GO:0022617; Response to fluid shear stress: GO:0034405; Leukocyte migration: GO:0050900.
